# Feeling Better Before, Not After: An Ecological Momentary Assessment of Affect Around Exercise in Women with Eating Disorders

**DOI:** 10.1101/2025.11.17.25340093

**Authors:** Danielle A. N. Chapa, Kelsie T. Forbush, Yiyang Chen, Courtney E. Costain, Samiya I. Rasheed

**Author notes:** CRediT author statement **Danielle Chapa:** Conceptualization, Data Curation, Formal Analysis (lead), Funding Acquisition, Investigation, Methodology, Project Administration, Writing-Original Draft Preparation (lead) **Kelsie Forbush:** Conceptualization, Resources, Supervision, Writing-Reviewing & Editing **Yiyang Chen:** Formal Analysis (support), Writing-Reviewing & Editing **Courtney Costain:** Writing-Reviewing & Editing **Samiya Rasheed:** Writing-Reviewing & Editing. Correspondence concerning this article should be addressed to Danielle A. N. Chapa, Department of Psychiatry, University of Pittsburgh, Pittsburgh, PA, 15213. Additional study materials (e.g., study protocol, data) may be available upon request.

## Abstract

Maladaptive exercise (MalE) includes excessive, compulsive, or compensatory exercise and is a common eating-disorder (ED) symptom associated with increased severity, slower rates-of-recovery, and faster rates-of-relapse. Affect-regulation theories posit that MalE functions to reduce high negative affect (NA), although support for the affect-regulation model is mixed. Previous studies have not integrated ecological momentary assessment (EMA) with accelerometry or examined the affect-regulation model in individuals with EDs who frequently engage in MalE. The objective of this study was to examine trajectories of NA and positive affect (PA) through a 7-day EMA study combined with wrist-worn accelerometry in women with EDs (*N*=84). Piecewise generalized mixed-effects regression models evaluated the trajectories of PA and NA in the hours leading up to and following self-reported exercise and exercise identified via accelerometry. NA was increasing before self-reported exercise, although NA did not meaningfully change relative to objectively measured exercise. PA was increasing prior to exercise and decreasing after exercise, and this pattern was consistent for both self-reported and objectively measured exercise. Rates of rising PA were steeper in the hours leading up to higher intensity exercise episodes. Though inconsistent with affect-regulation models, the current study offers preliminary evidence that exercise is associated with disrupted affective responses among women with EDs who regularly engage in MalE. Results suggest that planning or anticipating high-intensity exercise may be rewarding for a considerable proportion of people with EDs. If replicated, treatments may consider decreasing the reward value placed on intense exercise and increasing value placed on low-intensity or non-exercise activities.

---

Exercise, defined as any physical activity undertaken to improve health or fitness, is associated with benefits to both physical and mental health (Chekroud et al., 2018; Reiner et al., 2013; Rosenbaum et al., 2014). People with eating disorders (EDs) often report engaging in exercise to improve health, for enjoyment, or as a coping strategy for difficult emotions (Lampe et al., 2021), and when incorporated appropriately into ED treatment (Cook et al., 2016), exercise can promote improvements in physical health (e.g., increased bone density through strength training) and mental well-being (Bratland-Sanda et al., 2012; Hall et al., 2016; Pacanowski et al., 2017). Although exercise may offer benefits for individuals with EDs, it can also become maladaptive when motivated by weight or shape control, used to compensate for eating, or performed in a compulsive manner (Dittmer et al., 2018; Holland et al., 2014; Meyer & Taranis, 2011; Noetel et al., 2016). Maladaptive exercise (MalE) is a broad term meant to capture the full spectrum of harmful exercise. As such, MalE may include exercise completed at extreme intensities or durations (i.e., excessive exercise), intended to compensate for loss-of-control (LOC) eating (i.e., compensatory exercise), or completed despite serious illness or injury or with extreme rigidity (i.e., compulsive exercise) (Adkins & Keel, 2005; Dalle Grave et al., 2008; Holland et al., 2014; Mond et al., 2004; Mond et al., 2006; Monell et al., 2018).

MalE is common among individuals with EDs and frequently necessitates intervention due to its link with poor treatment prognosis. MalE is a transdiagnostic ED symptom, present in approximately 80% of individuals with anorexia nervosa (AN), 73% with atypical AN, and 40% with bulimia nervosa (BN) (Gorrell et al., 2021). Individuals who regularly engage in MalE report lower quality of life and higher levels of depression, anxiety, obsessive thoughts, and compulsive behaviors (Peñas-Lledó et al., 2002; Shroff et al., 2006; Young et al., 2018). Furthermore, MalE interferes with treatment and recovery, being associated with greater ED severity, lower remission rates, longer hospitalizations, and faster relapse (Dalle Grave et al., 2008; Monell et al., 2018; Solenberger, 2001; Stiles-Shields et al., 2015; Strober et al., 1997). Treatments targeting MalE are critical for improving outcomes and supporting long-term recovery, although the mechanisms underlying MalE remain insufficiently understood.

One promising direction for understanding the mechanisms that maintain MalE involves examining the role of affect (Meyer & Taranis, 2011), given that (1) exercise is known to modulate affect in the general population (Kanning & Schlicht, 2010; Liao et al., 2017), and (2) affect-regulation models of ED behaviors, such as binge eating and purging, have empirical support (Haedt-Matt & Keel, 2015; Schaefer et al., 2020; Wonderlich et al., 2022). The affect-regulation model of MalE posits that increasing negative affect (NA) triggers MalE, which functions to reduce NA. Ecological momentary assessment (EMA) study designs are well suited to identify trajectories of affect leading up to and following a behavior of interest in a naturalistic environment. Two prior studies examined trajectories of affect before and after exercise among individuals with EDs (Engel et al., 2013; Lampe et al., 2023). Contrary to the affect-regulation model, elevations in PA, not NA, preceded exercise (Engel et al., 2013; Lampe et al., 2023). PA increased in the hours following adaptive exercise; however, PA decreased in the hours following MalE (Lampe et al., 2023). Together, the existing literature suggests that exercise may be less of a reactive behavior to rising NA and more of a highly anticipated planned activity that is associated with increasing PA in the hours prior to the activity. Even though exercise is highly anticipated, its effects on PA may vary depending on whether the activity was adaptive or maladaptive.

Two key limitations exist in our current understanding of affective antecedents and consequences of MalE. First, individuals who do and do not report MalE in the months preceding naturalistic observation have often been combined resulting in heterogeneous samples. Evidence from non-ED samples found that pre-existing tendencies toward MalE moderated affect change, such that individuals with higher MalE at baseline showed steeper increases in PA prior to exercise relative to individuals with lower levels of MalE (Harris et al., 2025). These findings suggest that affective responses surrounding exercise may be different for individuals with established and routine MalE tendencies. Second, prior studies have rarely integrated accelerometer data into EMA designs. The use of accelerometers could enhance precision in identifying exercise onset and offset. Trying to approximate time of exercise in EMA surveys is limited because it relies on participants to initiate an event-contingent survey, accurately report exercise time retrospectively, or provides only a range of time that exercise occurred (i.e., between two surveys). Incorporation of accelerometers into EMA studies would also allow examination of how exercise intensity and duration influence affective change. This is particularly relevant given prior evidence that moderate doses of exercise (e.g., 30 minutes of moderate activity) increased PA (Hansen et al., 2001), whereas vigorous or prolonged exercise is associated with decreased PA (Reed & Ones, 2001). Understanding how dose of exercise (i.e., duration and intensity) shapes affective trajectories could have important clinical implications for tailoring ED treatment approaches in individuals with a historic pattern of MalE.

The overall objective of this study was to replicate prior research on the affect-regulation model of MalE using an EMA design, while extending previous work by sampling only women with EDs who engage in regular MalE and by combining self-reported exercise with wrist-worn accelerometry to examine exercise dose as a moderator of affective trajectories. We aimed to characterize trajectories of NA and PA leading up to and following: 1) self-reported and 2) accelerometer-based exercise. Consistent with the affect-regulation model, we hypothesized that increasing NA would precede exercise and affect would improve following exercise (i.e., increasing PA and decreasing NA). We also hypothesized that exercise dose would moderate post-exercise affect trajectories.

## Method

### Participants

Participants (*N*=84) were recruited from an active registry of community members with EDs and through study flyers posted around the community and on social media (see **Figure 1** for recruitment details). Interested participants completed an online screening questionnaire (details available in Supplemental Materials). Study inclusion criteria were: 1) current *DSM-5* ED; 2) age of 18 to 65 years; 3) frequent MalE in prior three months; 4) female sex; 5) fluency in written and spoken English; and 6) access to a mobile smartphone. Frequent MalE was defined as scoring one standard deviation higher than ED outpatients (Coniglio et al., 2018) on a measure of MalE, the Eating Pathology Symptoms Inventory (EPSI) Excessive Exercise scale (Forbush et al., 2013), or reporting MalE three times per week, on average, in the past three months on an adapted version of the Eating Disorder Diagnostic Scale for DSM-5 that assessed compulsive and compensatory exercise episodes (additional details in Supplemental Materials). Study exclusion criteria were: 1) medical conditions that affected appetite or body weight (e.g., untreated thyroid disorder, diabetes, current pregnancy); 2) medications that affected appetite or body weight (e.g., corticosteroids, stimulants); 3) current psychiatric diagnosis that could interfere with EMA protocol adherence (e.g., neurological disorder, current psychosis, and/or current manic episode); 4) significant visual impairment that would interfere with viewing and completing EMA surveys; 5) chronic or acute health problems that limited physical activity (e.g., broken leg, sprained ankle, pulled muscles, cardiopulmonary conditions); and 6) Division 1 collegiate athlete status due to high levels of sport-related activity that may have differential effects on affective trajectories.

**Figure 1.**
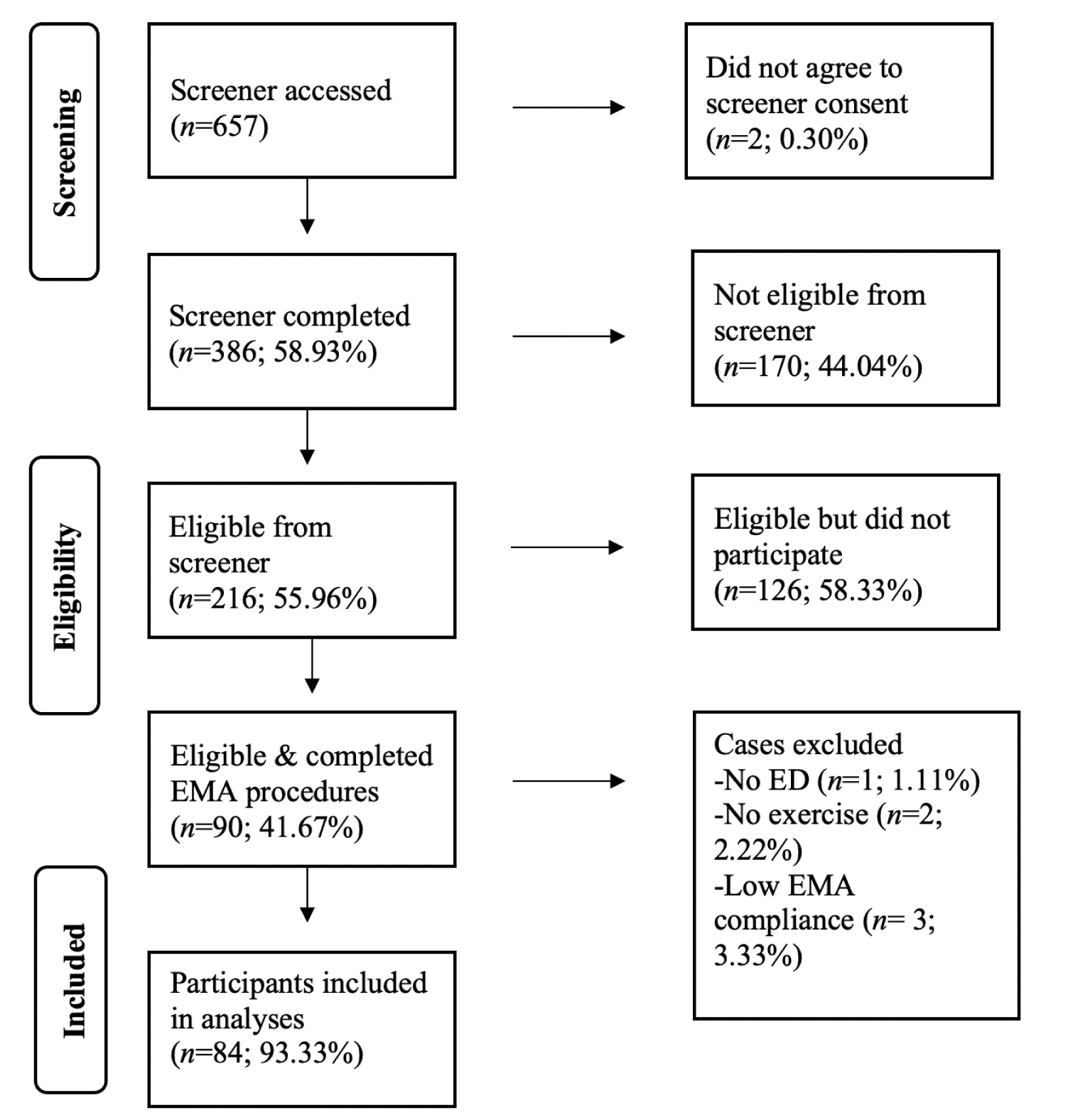
Study Recruitment.

### Procedure

The University of Kansas Institutional Review Board approved study procedures. Baseline appointments were scheduled for 90 minutes, either in-person or virtually. Participants completed two semi-structured interviews to confirm ED diagnosis and MalE behaviors (see Supplemental Materials for details). Participants downloaded the PiLR Health EMA App on their personal smartphone and completed a brief training on EMA study procedures (e.g., instructions on using the accelerometer and responding to surveys).

During the 7-day EMA protocol, participants were instructed to: 1) wear a research-grade accelerometer, the CentrePoint® Insight Watch, on their non-dominant wrist 24-hour per day; 2) respond daily to six, signal-contingent surveys on momentary PA and NA; and 3) complete event-contingent assessments of PA and NA before and after exercise. Participants were signaled six times per day at semi-random intervals, approximately every two to three hours, to complete a survey on momentary affect and eating behaviors. Signaled surveys expired after 30 minutes, and reminder notifications were sent 15 minutes prior to the expiration. To limit reactivity effects and any adverse effects associated with self-monitoring, participants did not have access to their physical activity data on the accelerometer screen (see Supplemental Materials for additional EMA details). Participants earned $2.00 each day they wore the accelerometer for at least 12-waking hours. If participants completed 34 or more assessments of affect, either signal- or event-contingent, they received an additional $36.00 for maximum compensation of $50.00.

### EMA Measures

#### Positive and Negative Affect Schedule (PANAS)

The PANAS (Watson et al., 1988) is comprised of twenty items that assess general PA and NA. To limit participant burden and increase EMA adherence, an adapted, shorter version of the PANAS was used in the current study with permission from the author (D. Watson, personal communication, 2/15/2019). The original PANAS was reduced from 20 items to ten items for the current study. Selected PA items included: interested, enthusiastic, determined, strong, and inspired. Selected NA items included: scared, nervous, distressed, ashamed, and irritable. The adapted version of the PANAS was administered to assess momentary PA and NA. The PANAS demonstrated evidence for excellent internal consistency when measuring momentary affect (Watson et al., 1988) (PA ω = .91 and NA ω = .82 in the current study). The PANAS has shown evidence for excellent convergent validity with other measures of PA and NA (Watson et al., 1988).

#### CentrePoint® Insight Watch (CP Insight)

The CP Insight Watch is a research-grade, wrist-worn accelerometer manufactured by Ametris (previously ActiGraph). During the 7-day EMA study period, participants were instructed to wear the CP Insight watch on the wrist of their non-dominant hand for 24 hours per day, except when their wrist would be submerged under water for extended periods. The CP Insight Watches were added to the protocol after the first ten participants completed the study, and two participants’ data were not available due to technical errors when initializing the device. CP Insight Watches recorded raw acceleration data using a triaxial accelerometer that quantifies movement in three different planes or axes, and raw data are summarized into activity counts (i.e., value indicating the intensity of movement/physical activity). The CP Insight watch was initialized according to standardized procedures (see Supplemental Materials).

### Statistical Approach

This study aimed to characterize trajectories of NA and PA leading up to and following: 1) self-reported and 2) accelerometer-based exercise. Self-reported exercise was identified as the time that an exercise event-contingent survey was submitted. When self-reported exercise occurred more than once in a day, only the first exercise event was used and only the affect ratings before the second event were modeled to avoid conflating two different exercise events. Accelerometer-based exercise time was identified as the start time of the longest moderate-to-vigorous physical activity (MVPA) episode derived from the CP Insight watch (see data processing details in Supplemental Materials).

Multilevel modeling (MLM) techniques were implemented in R Version 4.4.1. The lme() function within the nlme package (Pinheiro et al., 2025) was used to build a piecewise generalized mixed-effects regression model consistent with recommendations for testing affect-regulation models in EDs (Berg et al., 2013). Momentary affective observations (level 1) were nested within individual participants (level 2). Within-person residuals were modeled with a continuous autoregressive correlation structure to account for the impact of time and unequal distances between assessment time points. Time of exercise was mean centered at zero, and piecewise linear and quadratic functions of standardized time relative to exercise (i.e., hours prior to exercise and hours following exercise) predicted log transformed PA and NA. Log transformations were completed due to positive skewness of PA and NA ratings. Following best practices for building MLM models to analyze EMA data (Schwartz & Stone, 1998), a series of nested models were constructed incrementally adding polynomial linear and quadratic terms, among which we selected the best-fitting model based on model comparison results. See **Table 1** for nested model details. Slopes of the best-fitting models were used for hypothesis testing. *T*-tests for the pre-exercise trajectories test if slope estimates are significantly different from zero. *T*-tests for the post-exercise trajectories test if slope estimates are significantly different from the pre-exercise trajectory representing a significant change in direction or acceleration of affect. Additional analyses were completed to test if the post-exercise slope estimates were significantly different from zero by fitting a different model with alternative parameterization. Finally, calories burned was entered as a continuous moderator of objectively measured exercise to examine the extent that exercise dose impacted the slope of affect in the hours prior to and following exercise. Calories burned was selected as a proxy for exercise dose as calories burned is a function of movement intensity and duration. See Supplemental Materials for additional analytic details, R code, and R output.

**Table 1.** Fixed and Random Effects Tested in Nested Models.

| Model | Fixed Effects | Random Effects |
| --- | --- | --- |
| Model A |  | Random ID Intercept |
| Model B | (exerciseHours)<br>(exerciseHours:postExercise) | Random ID Intercept |
| Model C | (exerciseHours)<br>(exerciseHours:postExercise)<br>(exerciseHours)<sup>2</sup><br>(exerciseHours:postExercise)<sup>2</sup> | Random ID Intercept |
| Model D | (exerciseHours)<br>(exerciseHours:postExercise)<br>(exerciseHours)<sup>2</sup><br>(exerciseHours:postExercise)<sup>2</sup><br>(exerciseHours)<sup>3</sup><br>(exerciseHours:postExercise)<sup>3</sup> | Random ID Intercept |

*Note.* An ‘exerciseHours’ variable was created to represent the number of standardized decimal hours; negative values represented the number of hours prior to exercise, and positive values represented the number of hours after exercise. A ‘postExercise’ dummy coded variable was created; ‘0’ values were assigned to affect ratings made in the hours prior to exercise, and ‘1’ values were assigned to affect ratings made in the hours after exercise. The ‘exerciseHours’ and ‘exerciseHours:postExercise’ effects were entered as predictors to model the pre-exercise and post-exercise trajectories of affect respectively. Squared values represent quadratic effects. After the best fitting model selected with fixed effects, additional random effects (beyond ID) were added sequentially in the following order: exerciseHours, exerciseHours:postexercise, exerciseHours2, exerciseHours2:postexercise, etc.

## Results

A total of *N*=90 participants were consented into the study and attended the baseline appointment. There were *n*=3 participants who were not eligible following the baseline interviews and were excluded from the study. An additional *n*=3 participants were excluded from analyses due to low EMA compliance (i.e., less than 50%). Thus, the final sample used for analyses was *N*=84. The mean (*SD*) age was 29.83 (*11.49*) years with a range of 18-64 years, and the mean BMI was 25.41 (*5.57*) with a range of 17.64-39.85. Most participants had a diagnosis of ‘Other Specified Feeding or Eating Disorder’ (69.05%; *n*=58) followed by BN (27.38%; *n*=23), AN (2.38% *n*=2), and binge eating disorder (BED) (1.19%, *n*=1). Most participants were White, Non-Hispanic, and college-educated (**Table 2**).

**Table 2.**
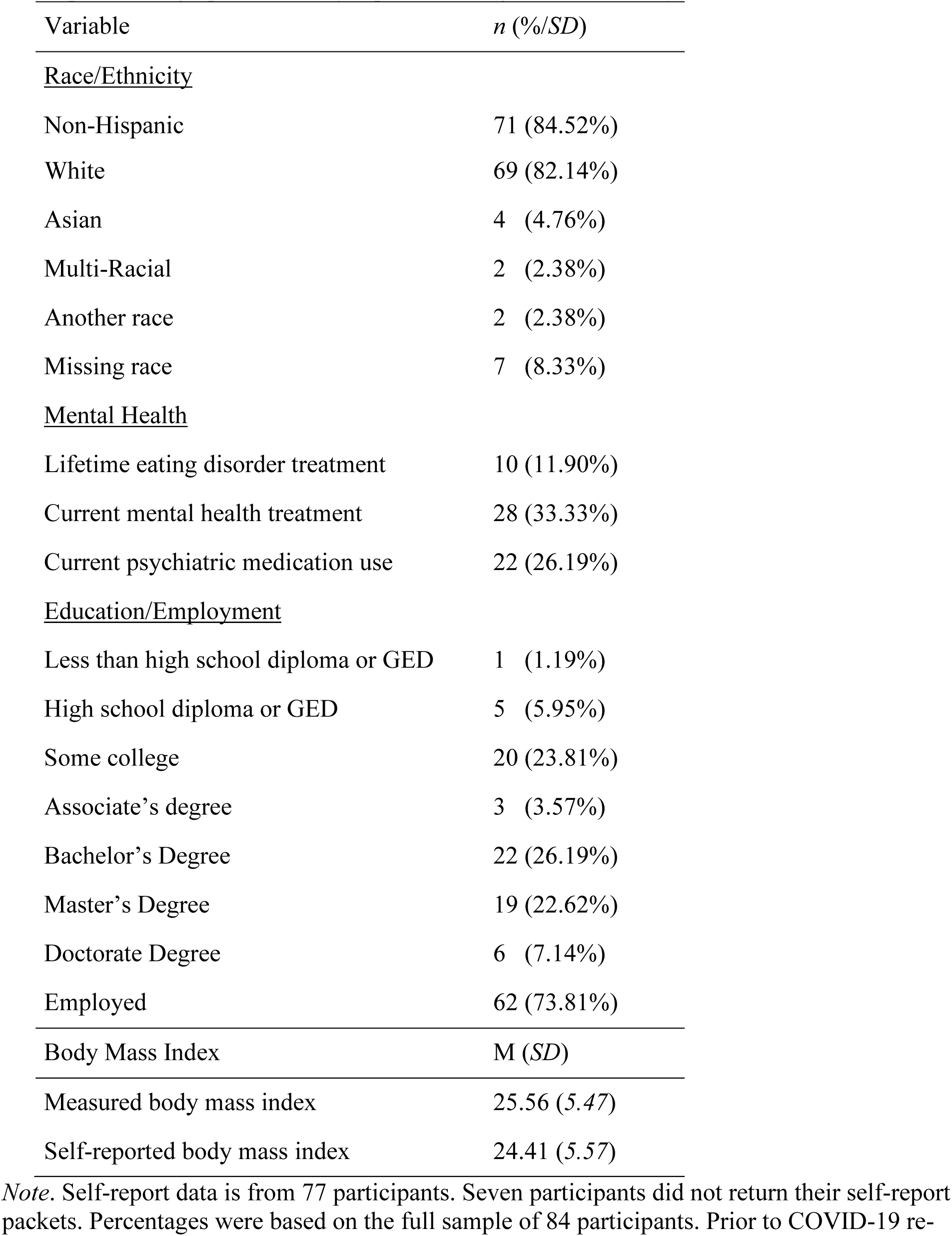
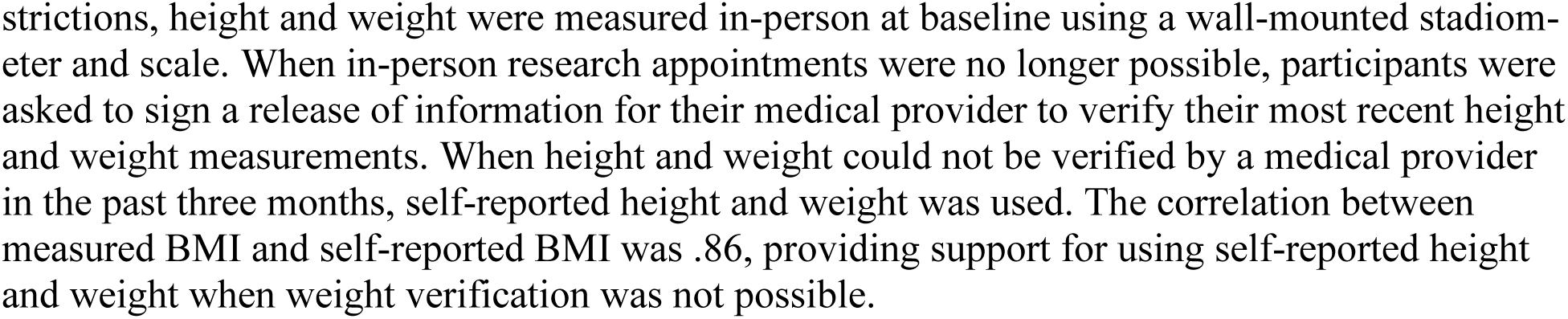
Participant demographics and self-reported eating-disorder history.

Participants completed an average of 34.4 signal-contingent surveys per week and an average of 7.15 event-contingent surveys per week. Participants were highly compliant with signal-contingent surveys (mean= 81.92%, median= 85.71%). Of the 72 individuals with accelerometer data available, participants had, on average, 6.5 days of valid accelerometer data (i.e., wear times of at least 12 waking hours).

### Self-Reported Exercise Results

Detailed model comparison results are available in the Supplemental Materials. Model C, with two fixed linear and two fixed quadratic slopes, emerged as the best fitting model for PA and NA. The addition of random effects for the pre- and post-exercise slopes did not improve model fit. PA increased curvilinearly in the hours leading up to exercise (linear estimate=0.013, quadratic estimate=0.077; **Table 3**) indicating that the rate of increase in PA accelerated in the hours closest to exercise (**Figure 2**). Following exercise, the linear slope of PA significantly deflected down (linear deflection=-0.309; **Table 3**) and PA decreased curvilinearly in the hours following exercise (linear estimate=-0.182, *SE*=0.029, *t*=-6.241, *p*<0.001; quadratic estimate=0.051, *SE*=0.020, *t*=2.501, *p*=0.013). In summary, PA increased in the hours prior to self-reported exercise and decreased in the hours following self-reported exercise.

**Figure 2.**
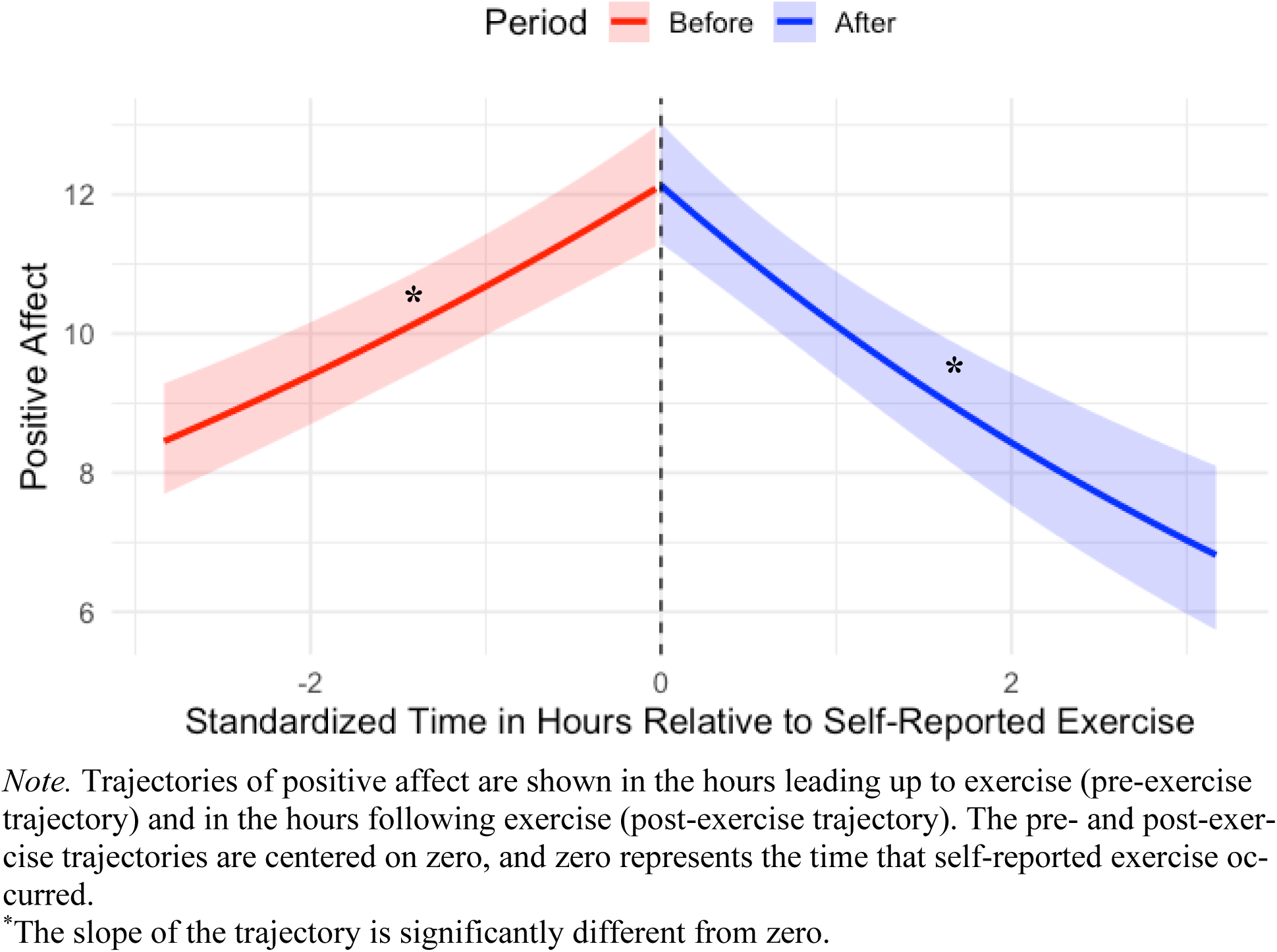
Positive Affect Trajectories Pre- and Post-Self-Reported Exercise.

**Table 3.** Multilevel Models for Positive and Negative Affective Relative to Self-Reported Exercise Time.

|  | Positive Affect |  |  | Negative Affect |  |  |
| --- | --- | --- | --- | --- | --- | --- |
|  | Est. | SE | t | Est. | SE | t |
| Intercept | 2.496 | 0.036 | 69.677*** | 1.931 | 0.032 | 60.523*** |
| Hours prior to event | 0.127 | 0.016 | 8.098*** | -0.078 | 0.014 | -5.535*** |
| (Hours prior to event)<sup>2</sup> | 0.077 | 0.017 | 4.647*** | -0.059 | 0.015 | -3.953*** |
| Hours following event | -0.309 | 0.041 | -7.467*** | 0.121 | 0.037 | 3.273** |
| (Hours following event)<sup>2</sup> | -0.027 | 0.019 | -1.426 | 0.041 | 0.017 | 2.416* |

*Note.* Est.= log-link estimates; SE= standard error. Fixed slope estimates for the pre-exercise and post-exercise trajectories are provided. Pre-exercise and post-exercise trajectories were centered on exercise time defined as the submission of an exercise event-contingent survey. The pre-exercise linear and quadratic slope estimates are represented by Hours prior to event and (Hours prior to event)<sup>2</sup> respectively. The post-exercise linear and quadratic slope estimates are represented by Hours following event and (Hours following event)<sup>2</sup> respectively. Significant *t*-tests for the pre-exercise trajectories suggest that the slope estimates are significantly different from zero. Significant *t*-tests for the post-exercise trajectories suggest that the slope estimates are significantly different from the pre-exercise trajectory. Additional analyses were completed to test if the post-exercise trajectory slopes were significantly different from zero. Results from those analyses found that the positive affect post-exercise trajectory linear and quadratic slopes were significantly different from zero. The negative affect linear and quadratic slope estimates for the post-exercise trajectory were both not significantly different from zero.
\* $p < .05$
\*\* $p < .01$
\*\*\* $p < .001$

NA decreased curvilinearly in the hours leading up to exercise (linear estimate=-0.078, quadratic estimate=-0.059; **Table 3**) indicating that the rate of decrease in NA accelerated in the hours closest to exercise (**Figure 3**). Following exercise, the linear and quadratic slopes of NA deflected upward (linear deflection=0.121, quadratic estimate=0.041; **Table 3**). NA stabilized following exercise (i.e., slope not significantly different from zero; linear estimate=0.043, *SE*=0.026, *t*=1.668, *p*=0.095; quadratic estimate=-0.018, *SE*=0.018, *t*=-0.993, *p*=0.321). In summary, NA declined in the hours prior to self-reported exercise and became constant after self-reported exercise.

**Figure 3.**
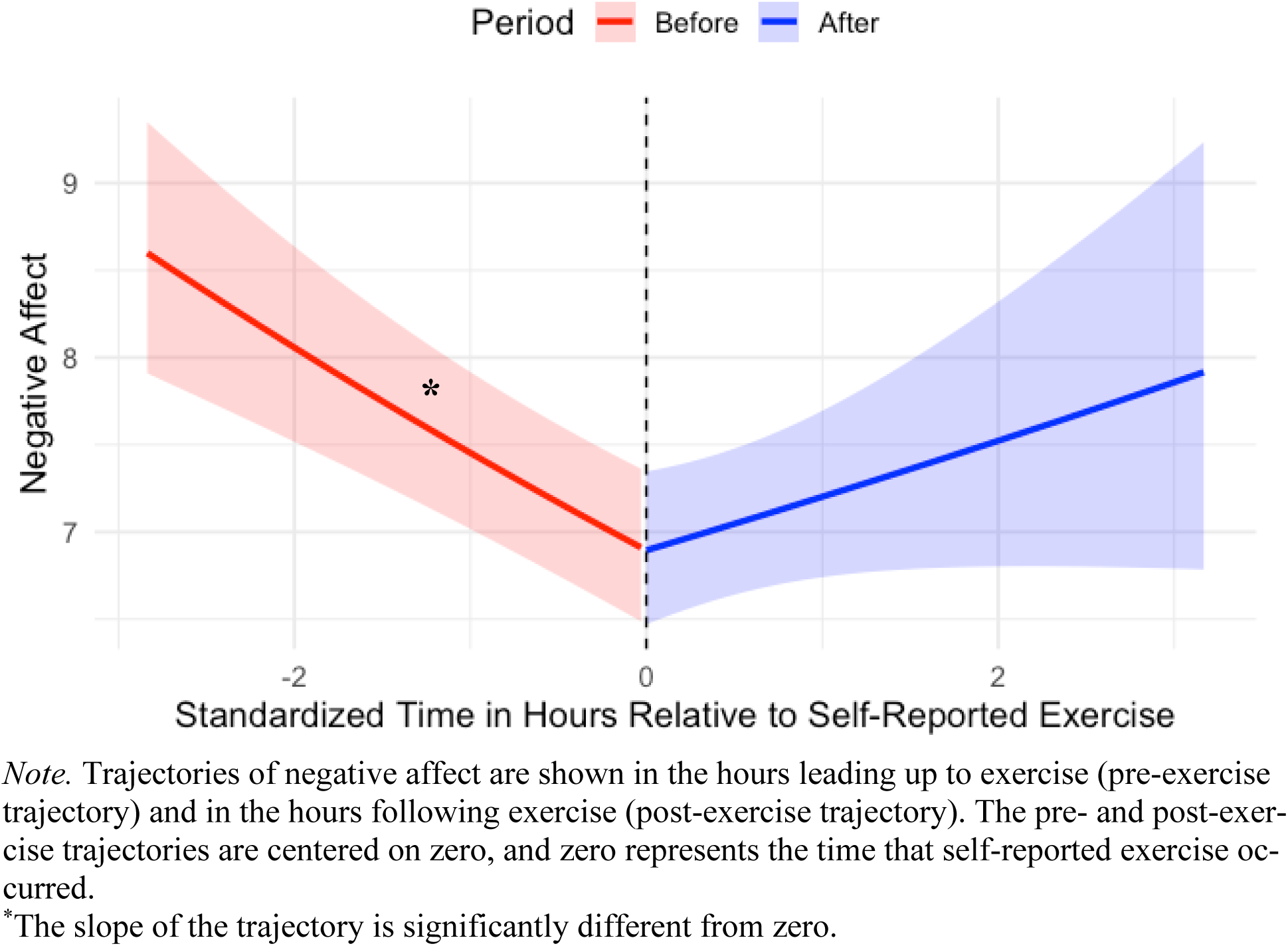
Negative Affect Trajectories Pre- and Post-Self-Reported Exercise.

### Objectively Measured Exercise Results

Detailed model comparison results are available in the Supplemental Materials. Model C, with two fixed linear and two fixed quadratic slopes, emerged as the best fitting fixed effects model for PA. Model A, with no fixed slopes and only a random intercept, emerged as the best fitting model for NA. PA increased curvilinearly in the hours leading up to exercise (linear estimate=0.134, quadratic estimate=0.111; **Table 4**). Thus, the rate of increase in PA accelerated in the hours closest to exercise (**Figure 4**). Following exercise, the trajectory of PA significantly deflected downward (linear deflection=-0.348; **Table 4**) and began decreasing (linear estimate=- 0.214, *SE*=0.039, *t*=-5.422, *p*<0.001; quadratic estimate=0.075, *SE*=0.025, *t*=3.060, *p*=0.002). In summary, PA increased in the hours prior to MVPA and decreased in the hours following MVPA. The NA slopes were not estimated because the best fitting model included only a random intercept indicating there was no significant change in NA prior to or following exercise. Caloric expenditure emerged as a significant moderator of pre-exercise change in PA, such that PA increased at a steeper rate in the hours prior to exercise that had higher caloric expenditure.

**Figure 4.**
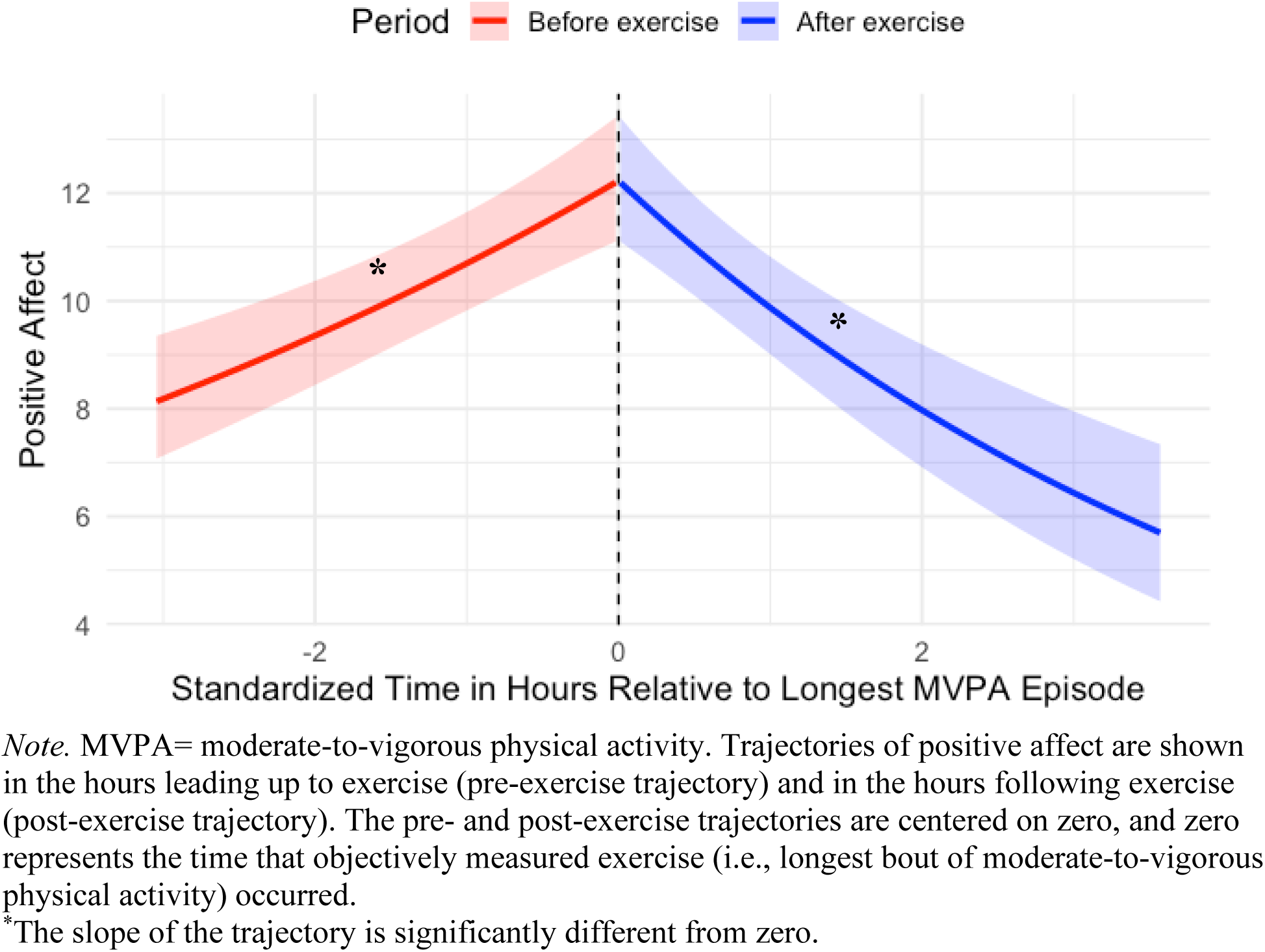
Positive Affect Trajectories Pre- and Post-Objectively Measured Exercise. *Note.* MVPA= moderate-to-vigorous physical activity. Trajectories of positive affect are shown in the hours leading up to exercise (pre-exercise trajectory) and in the hours following exercise (post-exercise trajectory). The pre- and post-exercise trajectories are centered on zero, and zero represents the time that objectively measured exercise (i.e., longest bout of moderate-to-vigorous physical activity) occurred. *The slope of the trajectory is significantly different from zero.

**Table 4.** Multilevel Models for Positive and Negative Affective Relative to Objectively Measured Exercise Time.

|  | Positive Affect |  |  | Negative Affect |  |  |
| --- | --- | --- | --- | --- | --- | --- |
|  | Est. | SE | *t* | Est. | SE | *t* |
| Intercept | 2.504 | 0.047 | 52.883*** | 2.026 | 0.036 | 56.071*** |
| Hours prior to event | 0.134 | 0.025 | 5.329*** | -- | -- | -- |
| (Hours prior to event)<sup>2</sup> | 0.111 | 0.024 | 4.608*** | -- | -- | -- |
| Hours following event | -0.348 | 0.061 | -5.696*** | -- | -- | -- |
| (Hours following event)<sup>2</sup> | -0.035 | 0.022 | -1.595 | -- | -- | -- |

*Note.* Est.= log-link estimates; SE= standard error. Fixed slope estimates for the pre-exercise and post-exercise trajectories are provided. Pre-exercise and post-exercise trajectories were centered on accelerometer-based physical activity time defined as the longest bout of moderate-to-vigorous activity. The pre-exercise linear and quadratic slope estimates are represented by Hours prior to event and (Hours prior to event)<sup>2</sup>. The post-exercise linear and quadratic slope estimates are represented by Hours following event and (Hours following event)<sup>2</sup>. Significant *t*-tests for the pre-exercise trajectories suggest that the slope estimates are significantly different from zero. Significant *t*-tests for the post-exercise trajectories suggest that the slope estimates are significantly different from the pre-exercise trajectory. Thus, in this table, the post-exercise trajectory estimates represent the amount of change or deflection in slope that was observed following exercise. Additional analyses were completed to test if the post-exercise trajectory slopes were significantly different from zero. Results from those analyses found that the positive affect post-exercise trajectory linear and quadratic slopes were significantly different from zero.
\* $p < .05$
\*\* $p < .01$
\*\*\* $p < .001$

## Discussion

The objective of this study was to examine trajectories of PA and NA leading up to and following exercise using both self-reported and accelerometer-based exercise. We hypothesized that increasing NA would precede exercise and decreasing NA would follow exercise consistent with the affect-regulation model. Contrary to our hypotheses, NA did not decrease following exercise. Rather, feelings of NA decreased in the hours leading up to self-reported exercise. This finding did not replicate when examining NA change prior to objectively identified MVPA. These inconsistencies may have emerged due to differences in a person’s first exercise episode of the day as opposed to their longest episode of MVPA. Overall, the affect-regulation model of MalE was not supported, highlighting that MalE may have a different function in individuals with an ED.

Anticipating or planning to engage in exercise appeared to improve mood, such that PA increased and NA decreased in the hours prior to exercise. Prior EMA studies also observed increases in PA occurring before exercise (Engel et al., 2013; Lampe et al., 2023). A risk and maintenance model of BN proposed that some ED behaviors transition from being impulsive during the early stages of an ED to compulsive as the ED progresses (Pearson et al., 2015). This theory could help contextualize the unexpected pre-exercise trajectory of increasing PA and decreasing NA observed in the current study. Many individuals with EDs engage in exercise to reduce NA (Bratland-Sanda et al., 2010; Noetel et al., 2016). Over time, NA may decline prior to exercise due to the anticipation/expectation that NA will decrease when exercise occurs. Exercise expectancies were not assessed in the current study, and future research is needed to test whether the relationships between MalE and momentary affect are mediated by expectations of affect change.

Another unexpected result was that PA decreased following exercise. Although not hypothesized, this finding is consistent with some prior EMA research (Engel et al., 2013; Lampe et al., 2023). For example, Lampe et al. (2023) found that MalE was associated with decreasing PA and adaptive exercise was associated with increasing PA. In the current study, decreases in PA were not moderated by exercise dose highlighting that the decline in PA following exercise was not a function of exercise intensity and duration. It is possible that decreased PA following exercise is related to distorted cognitions about exercise performance or the extent to which exercise goals were achieved (e.g., distorted evaluations about whether a workout was “good” or “bad” based on number of calories burned). Body checking or self-comparison during or immediately following exercise could similarly influence changes in affect (e.g., upward social comparisons during exercise).

Several limitations of the current study are important to discuss. First, the current study was limited to a sample of primarily White women, and therefore, the experiences of men and diverse races and ethnicities were not represented in the current study. Second, data collection of the current study overlapped with the COVID-19 pandemic. Thus, our results could have been influenced by the ongoing stressors and business closures associated with navigating the pandemic. For example, there was a period when gyms and other exercise facilities were closed. Qualitative and quantitative research on the effects of COVID-19 on EDs found that individuals reported exercising more overall, exercising more at home and outdoors, exercising more intensely, and feeling a higher drive to exercise (Brown et al., 2021; Schlegl et al., 2020; Termorshuizen et al., 2020). These studies suggest that MalE likely became more frequent during the COVID-19 pandemic. Third, individual exercise episodes during the EMA period where not distinguished as maladaptive vs. adaptive. The current study was designed to establish affective trajectories surrounding exercise among women with EDs who frequently engaged in MalE. However, it is possible that a portion of exercise episodes (identified by either self-report or accelerometer) could have been more adaptive in nature. Finally, potential limitations of the EMA design include possibility of reactivity effects associated with responding to prompts about mood and wearing an accelerometer. Many individuals with EDs already track their eating and physical activity using apps and passive sensors, which may limit the amount of reactivity effects associated with the EMA protocol. Additionally, some design features of the study were intended to limit reactivity effects, including one practice day and disenabling the function on the study accelerometer that displays exercise metrics to participants.

Despite limitations, the current study had several strengths. EMA procedures allowed for a longitudinal assessment of real-time changes in PA and NA relative to exercise. Physical activity and reports of affect were measured in free-living environments rather than in a laboratory or hospital setting, increasing ecological validity. Exercise time was evaluated using both self-reported initiation of exercise and MVPA identified with wrist-worn accelerometry. Self-reported exercise and wrist-worn accelerometry are associated with their own limitations and strengths. Self-reported exercise episodes require that participants remember to log an exercise episode in a timely manner, although one major strength is that it captures self-perceived exercise episodes regardless of duration or intensity of physical activity. The identification of exercise using an accelerometer is limited to the researcher-imposed definitions of what constitutes as an exercise episode. In the current study, our standardized definition of an MVPA episode could have excluded some types of exercise (e.g., traditional strength training with longer breaks or lower-intensity physical activity), although the addition of the accelerometer allowed us to assess the extent that exercise dose moderates affective trajectories. Finally, the current study also included a sample of individuals with a wide range of BMIs.

Although replication of the study results is needed, our results may have important implications for the treatment of MalE among individuals with EDs. Given that PA increased in the moments before exercise and decreased in the moments after exercise, treatment that focuses on helping people with EDs increase PA through the identification of enjoyable activities, other than exercise, could be helpful. Thus, scheduling positive activities using Behavioral Activation (Martell et al., 2021) could be beneficial for reducing MalE. Positive Affect Treatment (PAT), which is an adapted version of Behavioral Activation, was recently proposed to treat difficulties in reward processing among individuals with AN (Haynos et al., 2021) and may be efficacious for improving MalE as well. Results highlight that women with EDs who engage in regular MalE may not be receiving the affective benefits of exercise as demonstrated by decreased PA following exercise. Treatments aimed at helping individuals with EDs exercise in more adaptive ways should consider strategies to help people exercise in ways that improve their affect.

In conclusion, the current study used an EMA design combined with accelerometry to better understand exercise behaviors relative to momentary changes in affect. Preliminary longitudinal evidence suggests that exercise is associated with disrupted affective responses among women with EDs who regularly engage in MalE. Results consistently demonstrated that PA improved in the hours leading up to exercise, especially in the hours before intense exercise, as evidenced by increasing PA. In the hours after exercise, PA demonstrated a decreasing trajectory raising questions as to what interferes with the mood-boosting components of exercise for this population. Results suggest that planning or anticipating intense exercise episodes may be rewarding. If replicated, treatments may consider decreasing the reward value placed on high-intensity exercise and increasing the value placed on non-exercise activities to increase PA.

## Data Availability

Data produced in the present study may be available upon reasonable request to the authors.

