## Supplemental Materials for "Feeling Better Before, Not After: An Ecological Momentary Assessment of Affect Around Exercise in Women with Eating Disorders"

#### Section S1. Screening & Baseline Assessments

##### Screening Measures

###### *Eating Disorder Diagnostic Scale- DSM-5 Version (EDDS-5)*

The EDDS-5 was adapted from the original EDDS (Stice et al., 2000) which assessed *DSM-IV* ED symptoms. The EDDS-5 is a 22-item self-report diagnostic tool assessing *DSM-5* symptoms of AN, BN, BED, and OSFED in the previous three months. The EDDS-5 was used in the current study to screen for individuals who likely had an ED accompanied by high excessive exercise behaviors (i.e., at least three times per week, on average). The original EDDS includes one item assessing a specific component of maladaptive exercise, namely compensatory exercise (i.e., exercise intended to burn calories to compensate for overcompensation). Two additional screening items were added to assess other components of maladaptive exercise, How many times per week, on average, over the past 3 months have you "…engaged in exercise on a day when you were sick or injured?" and "…engaged in excessive exercise on a day when you were fasting (i.e., skipped 2 or more meals in a row)?". The EDDS demonstrated excellent criterion validity with interviewer-rated ED diagnoses, as well as excellent convergent validity with other measures of ED psychopathology (Stice et al., 2004b; Stice et al., 2000). The EDDS demonstrated excellent test-retest reliability (Stice et al., 2000) and internal consistency reliability (Stice et al., 2004b).

###### *Clinical Impairment Assessment (CIA)*

The CIA (Bohn et al., 2008) is a 16-item self-report of psychosocial impairment secondary to an ED over the past 28 days. The CIA assessed levels of ED-related clinical impairment at screening. Measuring clinical impairment was necessary to differentiate disordered eating from OSFED presentations. Without clinical impairment, self-reported ED symptoms that do not meet criteria for a *DSM-5* ED may be non-clinical disordered eating. Past research using receiver operator curves found that a score of 16 or greater on the CIA differentiates persons with a clinical ED from persons without an ED (Bohn et al., 2008). Thus, to be eligible for the current study, individuals with OSFED presentations were required to have a score of 16 or higher on the CIA. The CIA demonstrated excellent internal consistency as measured by Cronbach's alpha and item-total correlations (Bohn et al., 2008; Vannucci et al., 2012). The CIA also demonstrated acceptable test-retest reliability over three days and convergent validity with other measures of ED psychopathology (Bohn et al., 2008; Vannucci et al., 2012). Internal consistency reliability was excellent (a=.91) in the current study.

##### *Eating Pathology Symptoms Inventory (EPSI)*

The EPSI is a self-report measure of ED psychopathology (Forbush et al., 2013). The EPSI includes eight scales: Body Dissatisfaction, Binge Eating, Cognitive Restraint, Restricting, Excessive Exercise, Negative Attitudes Toward Obesity, and Muscle Building. In the current study, the EPSI was administered during screening to identify participants who reported high levels of excessive exercise, as indicated by a score of at least one standard deviation greater than normed scores of ED outpatients (Coniglio et al., 2018). The EPSI has shown evidence for a strong factor structure, excellent internal consistency, and excellent construct validity in past research (Coniglio et al., 2018; Forbush et al., 2013). In the current study, internal consistency reliability ranged from .60 for Cognitive Restraint to .93 for Negative Attitudes Toward Obesity.

##### *Additional Screening Items*

In addition to the EDDS and CIA, additional items were administered to assess remaining inclusion/exclusion criteria. These items assessed current age, English fluency, visual impairment, and ownership of a smartphone, as well as current medical diagnoses and medications that may affect appetite and body weight.

#### Baseline Measures

##### *Eating Disorder Diagnostic Interview (EDDI-5)*

The EDDI-5 (Presnell & Stice, 2003) is a clinician-rated interview that assesses *DSM-5* ED symptoms to generate ED diagnoses. The EDDI-5 was administered at baseline to confirm *DSM-5* ED diagnosis. In past studies, EDDI inter-rater reliability and test-retest reliability within one week were excellent (Stice et al., 2004, 2005, 2013).

##### Eating Pathology Symptoms Inventory- Clinician-Rated Version (EPSI-CRV)

The EPSI-CRV (Forbush et al., 2020) is a clinician-rated interview adapted from the EPSI self-report (Forbush et al., 2013). The EPSI-CRV includes thirteen modules to assess ED behaviors and cognitions. The Excessive Exercise Module of the EPSI-CRV measures maladaptive exercise behaviors (e.g., exercise despite being sick or injured) and beliefs (e.g., distress when unable to exercise and feeling compelled to exercise nearly every day) over the past three months. Detailed information about the type (e.g., running, recreational sports, lifting weights, etc.) and duration of exercise is collected within the EPSI-CRV Excessive Exercise Module. Exercise episodes are rated as to whether or not they meet diagnostic criteria for maladaptive exercise. The Excessive Exercise Module was administered at baseline to characterize exercise behaviors over the past three months. The EPSI-CRV Excessive Exercise Module demonstrated evidence for excellent convergent validity with the EPSI self-report Excessive Exercise scale and discriminant validity from a measure of anxiety and depression (Forbush et al., 2020). EPSI-CRV Excessive Exercise had good internal consistency as measured by Omega (Dunn et al., 2014) ($\omega$ =.71 in the current study).

#### Section S2. Study Procedures

Procedures were adapted during data collection due to COVID-19 regulations. All participants completed a baseline study visit, either in-person or virtually, to confirm ED diagnosis and to receive training on the 7-day EMA protocol. Prior to COVID-19, a subset of participants completed a second study visit following the EMA protocol to return study materials and receive payment. To limit face-to-face contact per COVID-19 regulations, the second study visit was suspended as of March 2020. Participants returned study materials and received reimbursement via USPS mail.

The default EMA schedule randomly triggered a survey within 30 minutes of the following time anchors: 9:00am, 11:30am, 2:00pm, 4:30pm, 7:00pm, and 9:30pm. The default schedule was adjusted to ensure that signal-contingent surveys were sent during waking hours and for individuals who were available only at the end of the hour (e.g., individuals whose jobs were based upon a 50-minute hour). To facilitate the completion of event-contingent surveys, participants could optionally create a geo-location for their typical exercise location (e.g., the address of their gym, park, indoor aquatics area, etc.). If a geo-location was created, participants received a reminder to complete the event-contingent exercise survey prior to exercise (i.e., when arriving to that location), and following exercise (i.e., when leaving that location).

#### Section S3. Accelerometer Procedures and Data Processing

Axis counts were collected in 60-second intervals or epochs. The wear criteria threshold was set to 720 minutes per day (i.e., 12-waking hours). Wear time validation settings included a minimum length of 60 minutes (i.e., number of minutes with consecutive zero counts to be considered non-wear) and a spike tolerance of 2 (i.e., consecutive number of epochs above the activity threshold needed before ending non-wear period). Wear compliance was calculated using an algorithm appropriate for adults (Toriano et. al., 2008).

The bouts() function within the Accelerometry package in R was used to identify all bouts of moderate-to-vigorous physical activity (MVPA) that were at least ten minutes in duration. Prior studies have not yet validated thresholds for identifying MVPA from activity counts derived from the CP Insight Watch. A threshold of 4,836 activity counts per minute was used in the current study to identify MVPA based on previous work validating activity count thresholds from a different ActiGraph wrist-worn triaxial accelerometer (Rhudy et al., 2020). To identify MVPA bouts, the activity count threshold was set to 4,836 activity counts (i.e., only minutes in which activity counts exceeded 4,836 were counted toward time in MVPA) with a tolerance of 2 minutes (i.e., no more than two minutes could include activity counts below 4,836). The longest activity bout was identified, and the start of each within-day longest activity bout was used as the objectively measured exercise time.

**Section S4. Statistical Analyses**

An ‘exerciseHours’ variable was created to represent the number of standardized decimal hours; negative values represented the number of hours prior to exercise, and positive values represented the number of hours after exercise. A ‘postExercise’ dummy coded variable was created; ‘0’ values were assigned to affect ratings made in the hours prior to exercise, and ‘1’ values were assigned to affect ratings made in the hours after exercise. The ‘exerciseHours’ and ‘exerciseHours:postExercise’ effects were entered as predictors to model the pre-exercise and post-exercise trajectories of affect separately.

Model comparisons were made at each incremental step to assess whether the addition of fixed linear and quadratic terms significantly improved model fit using -2 log likelihood ratio tests using the anova() function in R and evaluation of changes in Akaike’s Information Criteria (AIC) and Bayesian Information Criteria (BIC). Significant results from -2 log likelihood ratio tests suggested that the more complicated model significantly improved model fit with the addition of estimated parameters, in comparison with the relatively simpler model. Smaller AIC and BIC values also represented improved model fit. If the addition of model parameters did not contribute to significant model improvement or if the model would not converge, the parameters were dropped from the model. Following the selection of the best fitting model inclusive of fixed effects, random effects were similarly entered into the model incrementally.

A priori Power Analyses

With participants averaging three unhealthy exercise events per week and completing six signaled surveys of affect per day, we will have .80 power to detect an effect equivalent to Cohen's d=.54 for change in affect from pre to post-exercise episode with $N$=80 participants. A meta-analysis of 13 different EMA studies on EDs found that the weighted mean effect size for changes in affect pre and post ED symptoms (e.g., binge eating) was Cohen's d=.5 with a SD of .0773. Thus, our proposed study design is well powered to detect significant changes in affect

**Table S1**

*Positive Affect and Negative Affect Model Building Comparisons for Self-Reported Exercise Time- FIXED EFFECTS ONLY*

| Positive Affect Model Comparisons | | | | | | | |
|---|---|---|---|---|---|---|---|
| Model | *n* Parameters | AIC | BIC | -2LL | Residual *df* | $c^2$ | *df* |
| Model A | 4 | 703.763 | 726.441 | -347.882 | 2065 | -- | -- |
| Model B | 6 | 572.129 | 606.146 | -280.064 | 2063 | 135.635$^{***}$ | 2 |
| **Model C** | **8** | **555.170** | **600.526** | **-269.585** | **2061** | **20.959**$^{***}$ | **2** |
| Model D | 10 | 551.722 | 608.417 | -265.861 | 2059 | 7.448$^{*}$ | 2 |
| Negative Affect Model Comparisons | | | | | | | |
| Model | *n* Parameters | AIC | BIC | -2LL | Residual *df* | $c^2$ | *df* |
| Model A | 4 | 120.734 | 143.416 | -56.367 | 2067 | -- | -- |
| Model B | 6 | 98.264 | 132.287 | -43.132 | 2065 | 26.469$^{***}$ | 2 |
| **Model C** | **8** | **85.847** | **131.210** | **-34.923** | **2063** | **16.418**$^{***}$ | **2** |
| Model D | 10 | 80.815 | 137.519 | -30.408 | 2061 | 9.032$^{*}$ | 2 |

*Note.* AIC= Akaike information criterion; BIC= Bayesian information criterion; -2LL= -2 log likelihood test; *df*=degrees of freedom. Model comparison results are included for building the positive affect and negative affect models. Pre-exercise and post-exercise trajectories were centered on exercise time defined as the submission of an exercise event-contingent survey. Significant $c^2$ tests indicate that the model with additional parameters significantly improved model fit. Reduced AIC and BIC values also represent an improvement in model fit. Model a is the unconditional means model, which included only a random intercept. Model b added two fixed linear slopes (pre- and post-exercise trajectories). Model c added two fixed quadratic slopes. Model d added two fixed cubic slopes. $^{*}p < .05$ $^{**}$ $p < .01$ $^{***}p < .001$

**Table S2**

*Positive Affect and Negative Affect Model Building Comparisons for Objectively Measured Exercise Time-FIXED EFFECTS ONLY*

| Positive Affect Model Comparisons | | | | | | | |
|---|---|---|---|---|---|---|---|
| Model | *n* Parameters | AIC | BIC | -2LL | Residual | c$^2$ | *df* |
| Model a | 4 | 454.026 | 474.783 | -223.013 | 1264 | -- | -- |
| Model b | 6 | 420.940 | 452.075 | -204.470 | 1262 | 37.086$^{***}$ | 2 |
| **Model c** | **8** | **403.647** | **445.160** | **-193.823** | **1260** | **21.294**$^{***}$ | **2** |
| Model d | 10 | 401.048 | 452.940 | --190.524 | 1258 | 6.599$^{*}$ | 2 |
| Negative Affect Model Comparisons | | | | | | | |
| Model | *n* Parameters | AIC | BIC | -2LL | Residual | c$^2$ | *df* |
| **Model a** | **4** | **262.273** | **283.021** | **-127.137** | **1261** | **--** | **--** |
| Model b | 6 | 263.487 | 294.609 | -125.744 | 1259 | 2.786 | 2 |

*Note.* AIC= Akaike information criterion; BIC= Bayesian information criterion; -2LL= -2 log likelihood test; *df*=degrees of freedom. Model comparison results are included for building the positive and negative affect models. Pre-exercise and post-exercise trajectories were centered on accelerometer-based physical activity time defined as the longest bout of moderate-to-vigorous activity. Significant c$^2$ tests indicate that the model with additional parameters significantly improved model fit. Reduced AIC and BIC values also represent an improvement in model fit. Model a is the unconditional means model, which included only a random intercept. Model b added two fixed linear slopes (pre- and post-exercise trajectories). Model c added two fixed quadratic slopes. Model d added two fixed cubic slopes. $^{**}$ $p < .01$, $^{***}$ $p < .001$

### Self-Report Exercise Time

Danielle Chapa
11/13/25

The code below demonstrates models a-d using self-reported exercise time to model positive affect trajectories pre-exercise and post-exercise, including comparisons between models.

MODEL A: POSITIVE AFFECT

```
#MODEL A: baseline model containing a random effect for participant ID was cr
eated

PA_modelA <- lme(
  fixed = log_PosAff ~ 1,
  random = ~ 1 | ID,
  correlation = corCAR1(form = ~ StudyTime | ID),
  data = EMADat,
  na.action = na.omit,
  method = "ML"
)

summary(PA_modelA)

## Linear mixed-effects model fit by maximum likelihood
##   Data: EMADat
##        AIC      BIC    logLik
##   703.7633 726.4413 -347.8816
##
## Random effects:
##  Formula: ~1 | ID
##         (Intercept)  Residual
## StdDev:    0.280989 0.2815455
##
## Correlation Structure: Continuous AR(1)
##  Formula: ~StudyTime | ID
##  Parameter estimate(s):
##         Phi
## 0.006061274
## Fixed effects:  log_PosAff ~ 1
##                Value Std.Error   DF  t-value p-value
## (Intercept) 2.386399 0.0328848 2065 72.56846       0
##
## Standardized Within-Group Residuals:
##         Min          Q1         Med          Q3         Max
## -3.53194841 -0.61128352  0.05861564  0.70046066  3.52127770
##
## Number of Observations: 2142
## Number of Groups: 77
```

MODEL B: POSITIVE AFFECT

```
#MODEL B: two fixed linear slopes were added: 1) hours prior to exercise and
2) hours following exercise

PA_modelB <- lme(
  fixed = log_PosAff ~ 1 + exerciseHours_std + exerciseHours_std:dummytime,
  random = ~ 1 | ID,
  correlation = corCAR1(form = ~ StudyTime | ID),
  data = EMADat,
  na.action = na.omit,
  method = "ML"
)

summary(PA_modelB)

## Linear mixed-effects model fit by maximum likelihood
##   Data: EMADat
##        AIC      BIC    logLik
##   572.1286 606.1456 -280.0643
##
## Random effects:
##  Formula: ~1 | ID
##         (Intercept)  Residual
## StdDev:   0.2830692 0.2710888
##
## Correlation Structure: Continuous AR(1)
##  Formula: ~StudyTime | ID
##  Parameter estimate(s):
##         Phi
## 0.002623562
## Fixed effects:  log_PosAff ~ 1 + exerciseHours_std + exerciseHours_std:dum
mytime
##                                       Value  Std.Error   DF   t-value p-va
lue
## (Intercept)                       2.4539814 0.03357893 2063  73.08098
0
## exerciseHours_std                 0.0823780 0.01108956 2063   7.42843
0
## exerciseHours_std:dummytimeAfter -0.1894599 0.01653190 2063 -11.46026
0
##  Correlation:
##                                  (Intr) exrcH_
## exerciseHours_std                 0.141
## exerciseHours_std:dummytimeAfter -0.174 -0.810
##
## Standardized Within-Group Residuals:
##         Min          Q1         Med          Q3         Max
## -3.36674841 -0.60872997  0.06418156  0.69235011  3.73120125
##
```

```
## Number of Observations: 2142
## Number of Groups: 77
```

COMPARE A & B: POSITIVE AFFECT

```
#COMPARE A and B#
anova(PA_modelA, PA_modelB)

##           Model df      AIC      BIC    logLik   Test  L.Ratio p-value
## PA_modelA     1  4 703.7633 726.4413 -347.8816
## PA_modelB     2  6 572.1286 606.1456 -280.0643 1 vs 2 135.6346  <.0001
```

MODEL C: POSITIVE AFFECT

```
#MODEL C: two quadratic slopes were added

PA_modelC <- lme(
  fixed = log_PosAff ~ 1 + exerciseHours_std + exerciseHours_std:dummytime +
exerciseHours2_std + exerciseHours2_std:dummytime,
  random = ~ 1 | ID,
  correlation = corCAR1(form = ~ StudyTime | ID),
  data = EMADat,
  na.action = na.omit,
  method = "ML"
)
```

COMPARE B AND C: POSITIVE AFFECT

```
#COMPARE B and C#
anova(PA_modelB, PA_modelC)

##           Model df      AIC      BIC    logLik   Test  L.Ratio p-value
## PA_modelB     1  6 572.1286 606.1456 -280.0643
## PA_modelC     2  8 555.1699 600.5258 -269.5849 1 vs 2 20.95876  <.0001
```

MODEL D: POSITIVE AFFECT

```
#MODEL D: two fixed cubic slopes were added: 1) (hours prior to exercise)2 an
d 2) (hours following exercise)2

PA_modelD <- lme(
  fixed = log_PosAff ~ 1 + exerciseHours_std + exerciseHours2_std + exerciseH
ours_std:dummytime + exerciseHours2_std:dummytime + exerciseHours3_std + exer
ciseHours3_std:dummytime,
  random = ~ 1 | ID,
  correlation = corCAR1(form = ~ StudyTime | ID),
  data = EMADat,
  na.action = na.omit,
  method = "ML"
)

summary(PA_modelD)
```

```
## Linear mixed-effects model fit by maximum likelihood
##   Data: EMADat
##        AIC      BIC   logLik
##   551.722 608.4169 -265.861
##
## Random effects:
##  Formula: ~1 | ID
##         (Intercept)  Residual
## StdDev:   0.2839059 0.2686575
##
## Correlation Structure: Continuous AR(1)
##  Formula: ~StudyTime | ID
##  Parameter estimate(s):
##         Phi
## 0.001724259
## Fixed effects:  log_PosAff ~ 1 + exerciseHours_std + exerciseHours2_std +
exerciseHours_std:dummytime +      exerciseHours2_std:dummytime + exerciseHou
rs3_std + exerciseHours3_std:dummytime
##                                         Value  Std.Error   DF  t-value p-va
lue
## (Intercept)                          2.4136231 0.04991858 2059 48.35119  0.0
000
## exerciseHours_std                    0.1213803 0.03110344 2059  3.90247  0.0
001
## exerciseHours2_std                  -0.0125767 0.03739589 2059 -0.33631  0.7
367
## exerciseHours3_std                  -0.1176286 0.04710163 2059 -2.49734  0.0
126
## exerciseHours_std:dummytimeAfter   -0.1782475 0.08228569 2059 -2.16620  0.0
304
## exerciseHours2_std:dummytimeAfter -0.1441634 0.06193420 2059 -2.32769  0.0
200
## dummytimeAfter:exerciseHours3_std  0.2518852 0.09301753 2059  2.70793  0.0
068
##  Correlation:
##                                   (Intr) exrcH_ exrH2_ exrH3_ exH_:A eH2_:
A
## exerciseHours_std                  0.466
## exerciseHours2_std                 0.675  0.383
## exerciseHours3_std                 0.395 -0.299  0.694
## exerciseHours_std:dummytimeAfter  -0.710 -0.809 -0.776 -0.256
## exerciseHours2_std:dummytimeAfter  0.681  0.631  0.705  0.364 -0.890
## dummytimeAfter:exerciseHours3_std -0.646 -0.197 -0.892 -0.834  0.669 -0.78
9
##
## Standardized Within-Group Residuals:
##        Min         Q1        Med         Q3        Max
## -3.4253392 -0.5952212  0.0491465  0.6819538  3.7747835
##
```

```
## Number of Observations: 2142
## Number of Groups: 77
```

COMPARE C AND D: POSITIVE AFFECT

```
#COMPARE C and D#
anova(PA_modelC, PA_modelD)
```

```
##           Model df      AIC      BIC    logLik   Test  L.Ratio p-value
## PA_modelC     1  8 555.1699 600.5258 -269.5849
## PA_modelD     2 10 551.7220 608.4169 -265.8610 1 vs 2 7.447895  0.0241
```

The code below uses alternative dummy coding to test if the slopes for post-exercise positive affect are significantly different from zero.

```
#TEST IF POST-EXERCISE TRAJECTORY IS DIFFERENT THAN ZERO#
PA_post <- lme(
  fixed = log_PosAff ~ 1 + exerciseHours_std + exerciseHours2_std + exerciseH
ours_std:dummyReverse + exerciseHours2_std:dummyReverse,
  random = ~ 1 | ID,
  correlation = corCAR1(form = ~ StudyTime | ID),
  data = EMADat,
  na.action = na.omit,
  method = "ML"
)
```

The code below, creates a plot demonstrating change in positive affect pre- and post-exercise.

```
#set a range of possible X values (i.e., decimal exercise time)
grid_vals <- seq(
  min(EMADat$exerciseHours_std, na.rm = TRUE),
  max(EMADat$exerciseHours_std, na.rm = TRUE),
  length.out = 200
)

#using the final model, Model C, predict PA at each X value
emm <- emmeans(
  PA_modelC,
  specs = ~ exerciseHours_std | dummytime,
  at = list(exerciseHours_std = grid_vals)
)

#reverse log-transformed PA values
emm_df <- as.data.frame(emm)
emm_df <- emm_df %>%
  mutate(PA_pred = exp(emmean),
         PA_lower = exp(lower.CL),
         PA_upper = exp(upper.CL))
```

```r
# Split the data to model pre-exercise and post exercise
emm_before  <- subset(emm_df, dummytime == levels(emm_df$dummytime)[1] & exer
ciseHours_std <= 0)
emm_after   <- subset(emm_df, dummytime == levels(emm_df$dummytime)[2] & exer
ciseHours_std >= 0)

#create a visual plot
ggplot() +
  geom_ribbon(data = emm_before,
              aes(x = exerciseHours_std, ymin = PA_lower, ymax = PA_upper, fi
ll = dummytime),
              alpha = 0.2, color = NA) +
  geom_line(data = emm_before,
            aes(x = exerciseHours_std, y = PA_pred, color = dummytime),
            size = 1.2) +

  geom_ribbon(data = emm_after,
              aes(x = exerciseHours_std, ymin = PA_lower, ymax = PA_upper, fi
ll = dummytime),
              alpha = 0.2, color = NA) +
  geom_line(data = emm_after,
            aes(x = exerciseHours_std, y = PA_pred, color = dummytime),
            size = 1.2) +

  geom_vline(xintercept = 0, linetype = "dashed") +

  scale_color_manual(values = setNames(c("red", "blue"), levels(emm_df$dummyt
ime))) +
  scale_fill_manual(values  = setNames(c("red", "blue"), levels(emm_df$dummyt
ime))) +

  labs(
    x = "Standardized Time in Hours Relative to Self-Reported Exercise",
    y = "Positive Affect",
    color = "Period",
    fill = "Period",
    title = ""
  ) +
  theme_minimal(base_size = 14) +
  theme(legend.position = "top")

## Warning: Using `size` aesthetic for lines was deprecated in ggplot2 3.4.0.
## ℹ Please use `linewidth` instead.
## This warning is displayed once every 8 hours.
## Call `lifecycle::last_lifecycle_warnings()` to see where this warning was
## generated.
```

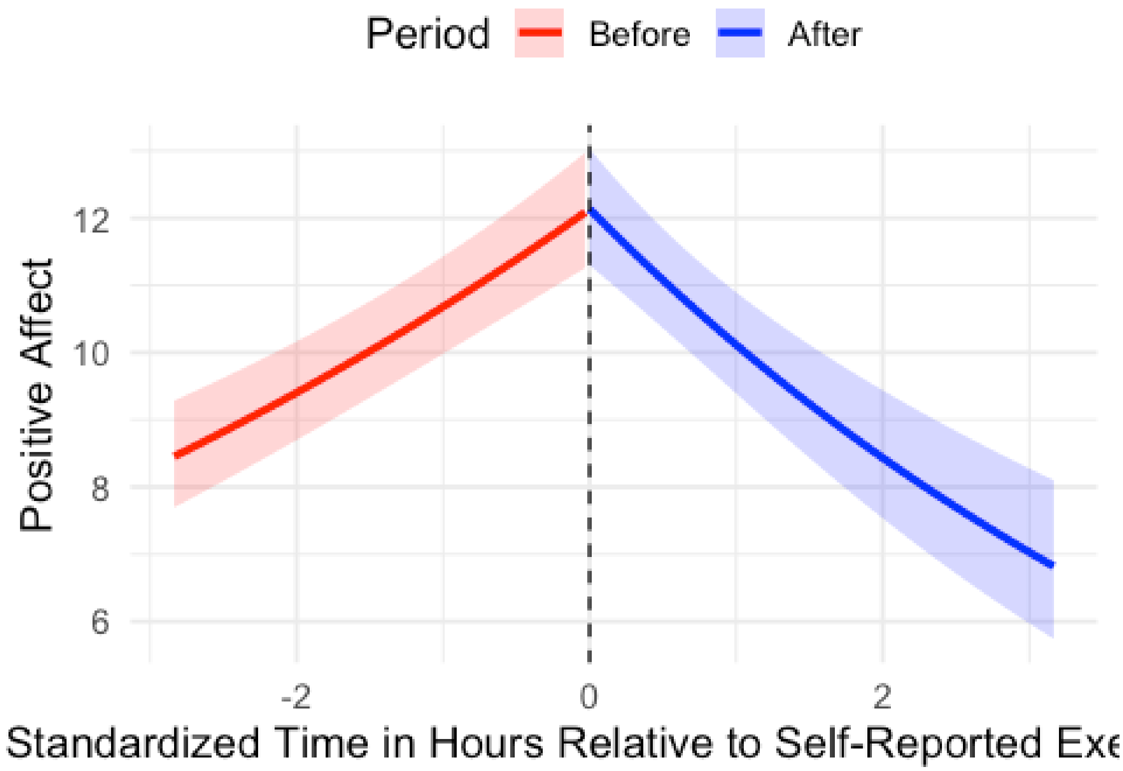


The code below demonstrates models a-d using self-reported exercise time to model negative affect trajectorie pre-exercise and post-exercise, including comparisons between models.

MODEL A: NEGATIVE AFFECT

```
#MODEL A: baseline model containing a random effect for participant ID was cr
eated
NA_modelA <- lme(
  fixed = log_NegAff ~ 1,
  random = ~ 1 | ID,
  correlation = corCAR1(form = ~ StudyTime | ID),
  data = EMADat,
  na.action = na.omit,
  method = "ML"
)
summary(NA_modelA)

## Linear mixed-effects model fit by maximum likelihood
##   Data: EMADat
##        AIC      BIC    logLik
##   120.734 143.4157 -56.36698
##
```

```
## Random effects:
##  Formula: ~1 | ID
##         (Intercept)  Residual
## StdDev:     0.25557 0.2447948
##
## Correlation Structure: Continuous AR(1)
##  Formula: ~StudyTime | ID
##  Parameter estimate(s):
##         Phi
## 0.004226975
## Fixed effects:  log_NegAff ~ 1
##                Value  Std.Error   DF  t-value p-value
## (Intercept) 1.975464 0.02984265 2067 66.19599       0
##
## Standardized Within-Group Residuals:
##        Min          Q1         Med          Q3         Max
## -3.5643224 -0.6837716 -0.1533759  0.5742621  3.8900201
##
## Number of Observations: 2144
## Number of Groups: 77
```

MODEL B: NEGATIVE AFFECT

```
#MODEL B: two fixed linear slopes were added: 1) hours prior to exercise and
2) hours following exercise
NA_modelB <- lme(
  fixed = log_NegAff ~ 1 + exerciseHours_std + exerciseHours_std:dummytime,
  random = ~ 1 | ID,
  correlation = corCAR1(form = ~ StudyTime | ID),
  data = EMADat,
  na.action = na.omit,
  method = "ML"
)
summary(NA_modelB)
```

```
## Linear mixed-effects model fit by maximum likelihood
##   Data: EMADat
##        AIC      BIC    logLik
##   98.26448 132.287 -43.13224
##
## Random effects:
##  Formula: ~1 | ID
##         (Intercept)  Residual
## StdDev:    0.252129 0.2429792
##
## Correlation Structure: Continuous AR(1)
##  Formula: ~StudyTime | ID
##  Parameter estimate(s):
##         Phi
## 0.003215456
```

```
## Fixed effects:  log_NegAff ~ 1 + exerciseHours_std + exerciseHours_std:dum
mytime
##                                       Value   Std.Error   DF  t-value p-va
lue
## (Intercept)                        1.9524891 0.029921589 2065 65.25352
0
## exerciseHours_std                 -0.0511160 0.009905821 2065 -5.16020
0
## exerciseHours_std:dummytimeAfter  0.0645773 0.014797788 2065  4.36399
0
##  Correlation:
##                                  (Intr) exrcH_
## exerciseHours_std                 0.142
## exerciseHours_std:dummytimeAfter -0.175 -0.809
##
## Standardized Within-Group Residuals:
##        Min         Q1        Med         Q3        Max
## -3.4398951 -0.6676346 -0.1292479  0.5740482  3.8655980
##
## Number of Observations: 2144
## Number of Groups: 77
```

COMPARE A AND B: NEGATIVE AFFECT

```
#COMPARE A and B#
anova(NA_modelA, NA_modelB)
```

```
##           Model df       AIC      BIC    logLik   Test  L.Ratio p-value
## NA_modelA     1  4 120.73397 143.4157 -56.36698
## NA_modelB     2  6  98.26448 132.2870 -43.13224 1 vs 2 26.46949  <.0001
```

MODEL C: NEGATIVE AFFECT

```
#MODEL C: two quadratic slopes were added
NA_modelC <- lme(
  fixed = log_NegAff ~ 1 + exerciseHours_std + exerciseHours_std:dummytime +
exerciseHours2_std + exerciseHours2_std:dummytime,
  random = ~ 1 | ID,
  correlation = corCAR1(form = ~ StudyTime | ID),
  data = EMADat,
  na.action = na.omit,
  method = "ML"
)
summary(NA_modelC)
```

```
## Linear mixed-effects model fit by maximum likelihood
##   Data: EMADat
##        AIC      BIC    logLik
##   85.84673 131.2102 -34.92336
##
## Random effects:
```

```
##  Formula: ~1 | ID
##         (Intercept)  Residual
## StdDev:   0.2526512 0.2413438
##
## Correlation Structure: Continuous AR(1)
##  Formula: ~StudyTime | ID
##  Parameter estimate(s):
##         Phi
## 0.001876358
## Fixed effects:  log_NegAff ~ 1 + exerciseHours_std + exerciseHours_std:dum
mytime +      exerciseHours2_std + exerciseHours2_std:dummytime
##                                        Value  Std.Error   DF  t-value p-va
lue
## (Intercept)                        1.9307445 0.03190092 2063 60.52316  0.0
000
## exerciseHours_std                 -0.0778821 0.01407014 2063 -5.53528  0.0
000
## exerciseHours2_std                -0.0585867 0.01482251 2063 -3.95255  0.0
001
## exerciseHours_std:dummytimeAfter   0.1214839 0.03711199 2063  3.27344  0.0
011
## dummytimeAfter:exerciseHours2_std  0.0405685 0.01679294 2063  2.41580  0.0
158
##  Correlation:
##                                   (Intr) exrcH_ exrH2_ exH_:A
## exerciseHours_std                  0.332
## exerciseHours2_std                 0.239  0.590
## exerciseHours_std:dummytimeAfter  -0.378 -0.854 -0.585
## dummytimeAfter:exerciseHours2_std  0.147  0.173 -0.346 -0.462
##
## Standardized Within-Group Residuals:
##        Min         Q1        Med         Q3        Max
## -3.4818815 -0.6648449 -0.1260921  0.5812239  3.8362967
##
## Number of Observations: 2144
## Number of Groups: 77
```

COMPARE B AND C: NEGATIVE AFFECT

```
#COMPARE B and C#
anova(NA_modelB, NA_modelC)
```

```
##           Model df      AIC      BIC    logLik   Test  L.Ratio p-value
## NA_modelB     1  6 98.26448 132.2870 -43.13224
## NA_modelC     2  8 85.84673 131.2102 -34.92336 1 vs 2 16.41775   3e-04
```

MODEL D: NEGATIVE AFFECT

```
NA_modelD <- lme(
  fixed = log_NegAff ~ 1 + exerciseHours_std + exerciseHours2_std + exerciseH
ours_std:dummytime + exerciseHours2_std:dummytime + exerciseHours3_std + exer
```

```
ciseHours3_std:dummytime,
  random = ~ 1| ID,
  correlation = corCAR1(form = ~ StudyTime | ID),
  data = EMADat,
  na.action = na.omit,
  method = "ML"
)
summary(NA_modelD)
```

```
## Linear mixed-effects model fit by maximum likelihood
##   Data: EMADat
##        AIC      BIC    logLik
##   80.81504 137.5193 -30.40752
##
## Random effects:
##  Formula: ~1 | ID
##         (Intercept)  Residual
## StdDev:   0.2535246 0.2410865
##
## Correlation Structure: Continuous AR(1)
##  Formula: ~StudyTime | ID
##  Parameter estimate(s):
##         Phi
## 0.002419862
## Fixed effects:  log_NegAff ~ 1 + exerciseHours_std + exerciseHours2_std +
exerciseHours_std:dummytime +      exerciseHours2_std:dummytime + exerciseHou
rs3_std + exerciseHours3_std:dummytime
##                                        Value  Std.Error   DF  t-value p-va
lue
## (Intercept)                        2.0151142 0.04469112 2061 45.08981  0.0
000
## exerciseHours_std                 -0.0666768 0.02790949 2061 -2.38904  0.0
170
## exerciseHours2_std                 0.0310008 0.03338723 2061  0.92852  0.3
532
## exerciseHours3_std                 0.1114583 0.04206280 2061  2.64981  0.0
081
## exerciseHours_std:dummytimeAfter  -0.0178225 0.07385790 2061 -0.24131  0.8
093
## exerciseHours2_std:dummytimeAfter  0.1649299 0.05567243 2061  2.96251  0.0
031
## dummytimeAfter:exerciseHours3_std -0.2506392 0.08333690 2061 -3.00754  0.0
027
##  Correlation:
##                                   (Intr) exrcH_ exrH2_ exrH3_ exH_:A eH2_:
A
## exerciseHours_std                  0.469
## exerciseHours2_std                 0.677  0.386
## exerciseHours3_std                 0.394 -0.299  0.692
## exerciseHours_std:dummytimeAfter  -0.712 -0.810 -0.777 -0.254
```

```
## exerciseHours2_std:dummytimeAfter  0.684  0.631  0.710  0.366 -0.891
## dummytimeAfter:exerciseHours3_std -0.647 -0.199 -0.892 -0.833  0.669 -0.79
2
##
## Standardized Within-Group Residuals:
##        Min         Q1        Med         Q3        Max
## -3.4105507 -0.6583342 -0.1219648  0.5898173  3.8153195
##
## Number of Observations: 2144
## Number of Groups: 77
```

COMPARAE C AND D: NEGATIVE AFFECT

```
anova(NA_modelC, NA_modelD)
```

```
##           Model df      AIC      BIC    logLik   Test  L.Ratio p-value
## NA_modelC     1  8 85.84673 131.2102 -34.92336
## NA_modelD     2 10 80.81504 137.5193 -30.40752 1 vs 2 9.031682  0.0109
```

The code below uses alternative dummy coding to test if the effects post-exercise negative affect are significantly different from zero.

```
#TEST IF POST-EXERCISE TRAJECTORY IS DIFFERENT THAN ZERO#

NA_post <- lme(
  fixed = log_NegAff ~ 1 + exerciseHours_std + exerciseHours_std:dummyReverse
+ exerciseHours2_std + exerciseHours2_std:dummyReverse,
  random = ~ 1 | ID,
  correlation = corCAR1(form = ~ StudyTime | ID),
  data = EMADat,
  na.action = na.omit,
  method = "ML"
)
summary(NA_post)
```

```
## Linear mixed-effects model fit by maximum likelihood
##   Data: EMADat
##        AIC      BIC    logLik
##   85.84673 131.2102 -34.92336
##
## Random effects:
##  Formula: ~1 | ID
##         (Intercept)  Residual
## StdDev:   0.2526512 0.2413438
##
## Correlation Structure: Continuous AR(1)
##  Formula: ~StudyTime | ID
##  Parameter estimate(s):
##         Phi
## 0.001876358
## Fixed effects:  log_NegAff ~ 1 + exerciseHours_std + exerciseHours_std:dum
```

```
myReverse +      exerciseHours2_std + exerciseHours2_std:dummyReverse
##                                     Value  Std.Error   DF  t-value p-valu
e
## (Intercept)                     1.9307445 0.03190092 2063 60.52316  0.000
0
## exerciseHours_std               0.0436018 0.02613640 2063  1.66824  0.095
4
## exerciseHours2_std             -0.0180182 0.01814901 2063 -0.99279  0.320
9
## exerciseHours_std:dummyReverse  -0.1214839 0.03711199 2063 -3.27344  0.001
1
## dummyReverse:exerciseHours2_std -0.0405685 0.01679294 2063 -2.41580  0.015
8
##  Correlation:
##                                 (Intr) exrcH_ exrH2_ exH_:R
## exerciseHours_std               -0.357
## exerciseHours2_std               0.331 -0.939
## exerciseHours_std:dummyReverse   0.378 -0.960  0.905
## dummyReverse:exerciseHours2_std -0.147  0.563 -0.643 -0.462
##
## Standardized Within-Group Residuals:
##        Min         Q1        Med         Q3        Max
## -3.4818815 -0.6648449 -0.1260921  0.5812239  3.8362967
##
## Number of Observations: 2144
## Number of Groups: 77
```

The code below, creates a plot demonstrating change in negative affect pre- and post-exercise.

```
#set a range of possible X values (i.e., decimal exercise time)
grid_vals <- seq(
  min(EMADat$exerciseHours_std, na.rm = TRUE),
  max(EMADat$exerciseHours_std, na.rm = TRUE),
  length.out = 200
)

#using the final model, Model C, predict NA at each X value
emmN <- emmeans(
  NA_modelC,
  specs = ~ exerciseHours_std | dummytime,
  at = list(exerciseHours_std = grid_vals)
)

#reverse log-transformed NA values
emmN_df <- as.data.frame(emmN)
emmN_df <- emmN_df %>%
  mutate(NA_pred = exp(emmean),
         NA_lower = exp(lower.CL),
         NA_upper = exp(upper.CL))
```

```
# Split the data to model pre-exercise and post exercise
emmN_before  <- subset(emmN_df, dummytime == levels(emmN_df$dummytime)[1] & e
xerciseHours_std <= 0)
emmN_after   <- subset(emmN_df, dummytime == levels(emmN_df$dummytime)[2] & e
xerciseHours_std >= 0)

#create a visual plot
ggplot() +
  geom_ribbon(data = emmN_before,
              aes(x = exerciseHours_std, ymin = NA_lower, ymax = NA_upper, fi
ll = dummytime),
              alpha = 0.2, color = NA) +
  geom_line(data = emmN_before,
            aes(x = exerciseHours_std, y = NA_pred, color = dummytime),
            size = 1.2) +

  geom_ribbon(data = emmN_after,
              aes(x = exerciseHours_std, ymin = NA_lower, ymax = NA_upper, fi
ll = dummytime),
              alpha = 0.2, color = NA) +
  geom_line(data = emmN_after,
            aes(x = exerciseHours_std, y = NA_pred, color = dummytime),
            size = 1.2) +

  geom_vline(xintercept = 0, linetype = "dashed") +

  scale_color_manual(values = setNames(c("red", "blue"), levels(emmN_df$dummy
time))) +
  scale_fill_manual(values  = setNames(c("red", "blue"), levels(emmN_df$dummy
time))) +

  labs(
    x = "Standardized Time in Hours Relative to Self-Reported Exercise",
    y = "Negative Affect",
    color = "Period",
    fill = "Period",
    title = ""
  ) +
  theme_minimal(base_size = 14) +
  theme(legend.position = "top")
```

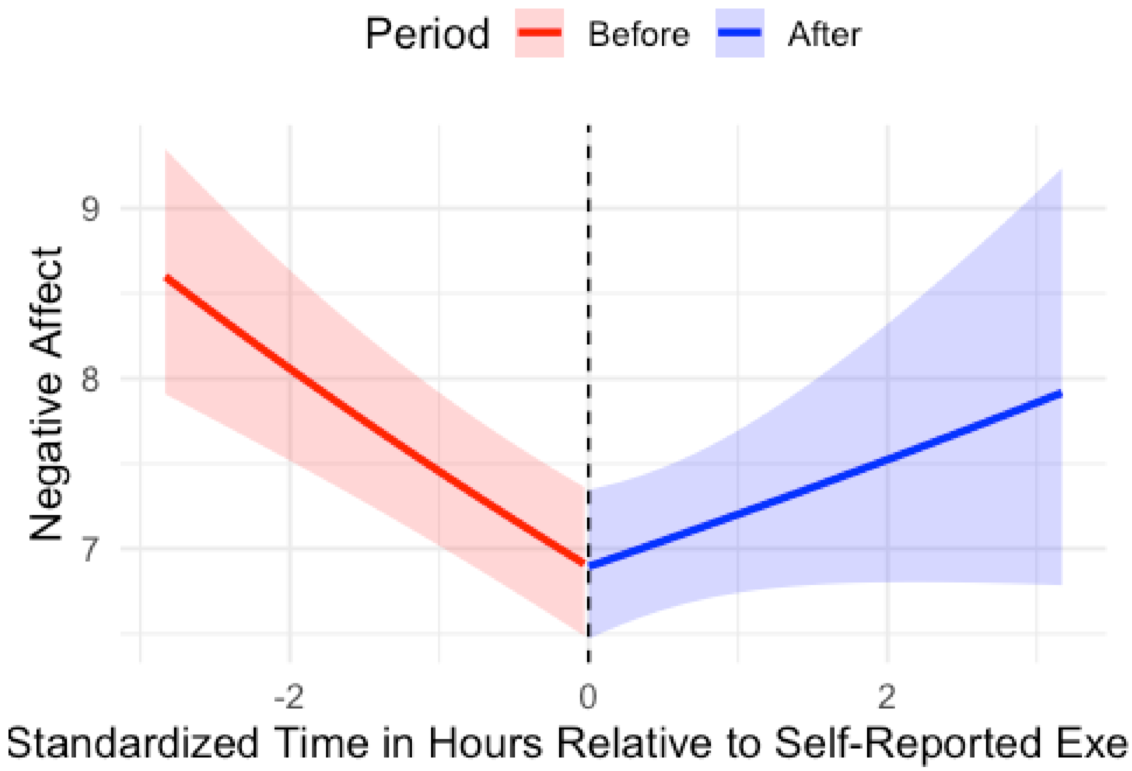
Period
Before
After
Negative Affect
9
8
7
-2
0
2
Standardized Time in Hours Relative to Self-Reported Exe

### Accelerometer-Based Exercise Time

Danielle Chapa

11/13/25

The code below demonstrates models a-d using ACCELEROMETER-BASED exercise time to model positive affect trajectories pre-exercise and post-exercise, including comparisons between models.

MODEL A: POSITIVE AFFECT

```
#MODEL A: baseline model containing a random effect for participant ID was cr
eated

PA_modelA <- lme(
  fixed = log_PosAff ~ 1,
  random = ~ 1 | ID,
  correlation = corCAR1(form = ~ StudyTime | ID),
  data = EMADat,
  na.action = na.omit,
  method = "ML"
)

summary(PA_modelA)

## Linear mixed-effects model fit by maximum likelihood
##   Data: EMADat
##        AIC      BIC    logLik
##   454.0263 474.783 -223.0132
##
## Random effects:
##  Formula: ~1 | ID
##         (Intercept) Residual
## StdDev:   0.3159183 0.276761
##
## Correlation Structure: Continuous AR(1)
##  Formula: ~StudyTime | ID
##  Parameter estimate(s):
##         Phi
## 0.006410992
## Fixed effects:  log_PosAff ~ 1
##                Value  Std.Error   DF t-value p-value
## (Intercept) 2.369331 0.04165638 1264  56.878       0
##
## Standardized Within-Group Residuals:
##         Min          Q1         Med          Q3         Max
## -3.62180374 -0.55834966  0.08135943  0.69175758  3.47801863
##
## Number of Observations: 1325
## Number of Groups: 61
```

MODEL B: POSITIVE AFFECT

```
#MODEL B: two fixed linear slopes were added: 1) hours prior to exercise and
2) hours following exercise

PA_modelB <- lme(
  fixed = log_PosAff ~ 1 + exerciseHours_std + exerciseHours_std:dummytime,
  random = ~ 1 | ID,
  correlation = corCAR1(form = ~ StudyTime | ID),
  data = EMADat,
  na.action = na.omit,
  method = "ML"
)

summary(PA_modelB)

## Linear mixed-effects model fit by maximum likelihood
##   Data: EMADat
##        AIC      BIC    logLik
##   420.9403 452.0754 -204.4702
##
## Random effects:
##  Formula: ~1 | ID
##         (Intercept) Residual
## StdDev:   0.3152858 0.272198
##
## Correlation Structure: Continuous AR(1)
##  Formula: ~StudyTime | ID
##  Parameter estimate(s):
##         Phi
## 0.004280982
## Fixed effects:  log_PosAff ~ 1 + exerciseHours_std + exerciseHours_std:dum
mytime
##                                              Value  Std.Error   DF  t-va
lue
## (Intercept)                              2.4221368 0.04268025 1262 56.75
076
## exerciseHours_std                        0.0485247 0.01550537 1262  3.12
954
## exerciseHours_std:dummytimeAfter exercise -0.1335955 0.02435176 1262 -5.48
607
##                                          p-value
## (Intercept)                               0.0000
## exerciseHours_std                         0.0018
## exerciseHours_std:dummytimeAfter exercise  0.0000
##  Correlation:
##                                          (Intr) exrcH_
## exerciseHours_std                         0.193
## exerciseHours_std:dummytimeAfter exercise -0.227 -0.842
##
```

```
## Standardized Within-Group Residuals:
##         Min          Q1         Med          Q3         Max
## -3.68462681 -0.57774917  0.06214749  0.66619531  3.67352003
##
## Number of Observations: 1325
## Number of Groups: 61
```

COMPARE A & B: POSITIVE AFFECT

```
#COMPARE A and B#
anova(PA_modelA, PA_modelB)

##           Model df      AIC      BIC    logLik   Test  L.Ratio p-value
## PA_modelA     1  4 454.0263 474.7830 -223.0132
## PA_modelB     2  6 420.9403 452.0754 -204.4702 1 vs 2 37.08598  <.0001
```

MODEL C: POSITIVE AFFECT

```
#MODEL C: two quadratic slopes were added
PA_modelC <- lme(
  fixed = log_PosAff ~ 1 + exerciseHours_std + exerciseHours_std:dummytime +
exerciseHours2_std + exerciseHours2_std:dummytime,
  random = ~ 1 | ID,
  correlation = corCAR1(form = ~ StudyTime | ID),
  data = EMADat,
  na.action = na.omit,
  method = "ML"
)

summary(PA_modelC)

## Linear mixed-effects model fit by maximum likelihood
##   Data: EMADat
##        AIC      BIC    logLik
##   403.6467 445.1601 -193.8234
##
## Random effects:
##  Formula: ~1 | ID
##         (Intercept)  Residual
## StdDev:   0.3154094 0.2696341
##
## Correlation Structure: Continuous AR(1)
##  Formula: ~StudyTime | ID
##  Parameter estimate(s):
##         Phi
## 0.003431966
## Fixed effects:  log_PosAff ~ 1 + exerciseHours_std + exerciseHours_std:dum
mytime +      exerciseHours2_std + exerciseHours2_std:dummytime
##                                                   Value  Std.Error   DF  t-v
alue
## (Intercept)                                   2.5037472 0.04734503 1260 52.8
```

```
8300
## exerciseHours_std                            0.1339285 0.02513297 1260  5.3
2880
## exerciseHours2_std                           0.1106496 0.02401042 1260  4.6
0840
## exerciseHours_std:dummytimeAfter exercise  -0.3478458 0.06106314 1260 -5.6
9649
## dummytimeAfter exercise:exerciseHours2_std -0.0353273 0.02214735 1260 -1.5
9510
##                                            p-value
## (Intercept)                                 0.0000
## exerciseHours_std                           0.0000
## exerciseHours2_std                          0.0000
## exerciseHours_std:dummytimeAfter exercise   0.0000
## dummytimeAfter exercise:exerciseHours2_std  0.1109
##  Correlation:
##                                            (Intr) exrcH_ exrH2_ eH_:Ae
## exerciseHours_std                           0.444
## exerciseHours2_std                          0.349  0.701
## exerciseHours_std:dummytimeAfter exercise  -0.477 -0.913 -0.704
## dummytimeAfter exercise:exerciseHours2_std  0.080  0.026 -0.434 -0.227
##
## Standardized Within-Group Residuals:
##         Min          Q1         Med          Q3         Max
## -3.64094413 -0.57238198  0.05694232  0.63047839  3.83709585
##
## Number of Observations: 1325
## Number of Groups: 61
```

COMPARE B AND C: POSITIVE AFFECT

```
#COMPARE B and C#
anova(PA_modelB, PA_modelC)

##           Model df      AIC      BIC    logLik   Test  L.Ratio p-value
## PA_modelB     1  6 420.9403 452.0754 -204.4702
## PA_modelC     2  8 403.6467 445.1601 -193.8234 1 vs 2 21.29361  <.0001
```

MODEL D: POSITIVE AFFECT

```
#MODEL D: two fixed linear slopes AND two fixed quadratic slopes AND two fixe
d cubic were added
PA_modelD <- lme(
  fixed = log_PosAff ~ 1 + exerciseHours_std + exerciseHours_std:dummytime +
exerciseHours2_std + exerciseHours2_std:dummytime + exerciseHours3_std + exer
ciseHours3_std:dummytime,
  random = ~ 1 | ID,
  correlation = corCAR1(form = ~ StudyTime | ID),
  data = EMADat,
  na.action = na.omit,
  method = "ML"
```

```
)
summary(PA_modelD)
```

```
## Linear mixed-effects model fit by maximum likelihood
##   Data: EMADat
##        AIC      BIC   logLik
##   401.0479 452.9396 -190.524
##
## Random effects:
##  Formula: ~1 | ID
##         (Intercept)  Residual
## StdDev:   0.3156791 0.2687843
##
## Correlation Structure: Continuous AR(1)
##  Formula: ~StudyTime | ID
##  Parameter estimate(s):
##         Phi
## 0.003062841
## Fixed effects:  log_PosAff ~ 1 + exerciseHours_std + exerciseHours_std:dum
mytime +      exerciseHours2_std + exerciseHours2_std:dummytime + exerciseHou
rs3_std +      exerciseHours3_std:dummytime
##                                                   Value  Std.Error   DF  t-v
alue
## (Intercept)                                   2.5943858 0.07274141 1258 35.6
6587
## exerciseHours_std                             0.1248522 0.04362641 1258  2.8
6185
## exerciseHours2_std                            0.2317660 0.06223717 1258  3.7
2392
## exerciseHours3_std                            0.1789355 0.07011653 1258  2.5
5197
## exerciseHours_std:dummytimeAfter exercise  -0.4631621 0.11949419 1258 -3.8
7602
## dummytimeAfter exercise:exerciseHours2_std -0.0053473 0.05906410 1258 -0.0
9053
## dummytimeAfter exercise:exerciseHours3_std -0.2629573 0.12091574 1258 -2.1
7472
##                                            p-value
## (Intercept)                                 0.0000
## exerciseHours_std                           0.0043
## exerciseHours2_std                          0.0002
## exerciseHours3_std                          0.0108
## exerciseHours_std:dummytimeAfter exercise   0.0001
## dummytimeAfter exercise:exerciseHours2_std  0.9279
## dummytimeAfter exercise:exerciseHours3_std  0.0298
##  Correlation:
##                                            (Intr) exrcH_ exrH2_ exrH3_ eH_
:Ae
## exerciseHours_std                           0.602
## exerciseHours2_std                          0.762  0.523
```

```
## exerciseHours3_std                                0.546  0.009  0.811
## exerciseHours_std:dummytimeAfter exercise  -0.792 -0.867 -0.831 -0.458
## dummytimeAfter exercise:exerciseHours2_std  0.644  0.728  0.565  0.296 -0.
819
## dummytimeAfter exercise:exerciseHours3_std -0.721 -0.364 -0.922 -0.900  0.
730
##                                            dAe:H2
## exerciseHours_std
## exerciseHours2_std
## exerciseHours3_std
## exerciseHours_std:dummytimeAfter exercise
## dummytimeAfter exercise:exerciseHours2_std
## dummytimeAfter exercise:exerciseHours3_std -0.649
##
## Standardized Within-Group Residuals:
##        Min         Q1        Med          Q3        Max
## -3.6445819 -0.5557512  0.0764637  0.6210919  3.8777018
##
## Number of Observations: 1325
## Number of Groups: 61
```

COMPARE C AND D: POSITIVE AFFECT

```
#COMPARE C and D#
anova(PA_modelC, PA_modelD)

##           Model df      AIC      BIC    logLik   Test L.Ratio p-value
## PA_modelC     1  8 403.6467 445.1601 -193.8234
## PA_modelD     2 10 401.0479 452.9396 -190.5240 1 vs 2 6.59881  0.0369
```

The code below uses alternative dummy coding to test if the slopes for post-exercise positive affect are significantly different from zero.

```
#TEST IF POST-EXERCISE TRAJECTORY IS DIFFERENT THAN ZERO#
PA_post <- lme(
  fixed = log_PosAff ~ 1 + exerciseHours_std + exerciseHours2_std + exerciseH
ours_std:dummyReverse + exerciseHours2_std:dummyReverse,
  random = ~ 1 | ID,
  correlation = corCAR1(form = ~ StudyTime | ID),
  data = EMADat,
  na.action = na.omit,
  method = "ML"
)
summary(PA_post)

## Linear mixed-effects model fit by maximum likelihood
##   Data: EMADat
##        AIC      BIC    logLik
##   403.6467 445.1601 -193.8234
##
## Random effects:
```

```
##  Formula: ~1 | ID
##         (Intercept)  Residual
## StdDev:   0.3154094 0.2696341
##
## Correlation Structure: Continuous AR(1)
##  Formula: ~StudyTime | ID
##  Parameter estimate(s):
##         Phi
## 0.003431966
## Fixed effects:  log_PosAff ~ 1 + exerciseHours_std + exerciseHours2_std +
exerciseHours_std:dummyReverse +      exerciseHours2_std:dummyReverse
##                                       Value  Std.Error   DF  t-value p-valu
e
## (Intercept)                       2.5037472 0.04734503 1260 52.88300  0.000
0
## exerciseHours_std                -0.2139173 0.03945682 1260 -5.42156  0.000
0
## exerciseHours2_std                0.0753223 0.02461383 1260  3.06016  0.002
3
## exerciseHours_std:dummyReverse    0.3478458 0.06106314 1260  5.69649  0.000
0
## exerciseHours2_std:dummyReverse   0.0353273 0.02214735 1260  1.59510  0.110
9
##  Correlation:
##                                  (Intr) exrcH_ exrH2_ exH_:R
## exerciseHours_std                -0.455
## exerciseHours2_std                0.412 -0.928
## exerciseHours_std:dummyReverse    0.477 -0.966  0.891
## exerciseHours2_std:dummyReverse -0.080  0.334 -0.477 -0.227
##
## Standardized Within-Group Residuals:
##         Min          Q1         Med          Q3         Max
## -3.64094413 -0.57238198  0.05694232  0.63047839  3.83709585
##
## Number of Observations: 1325
## Number of Groups: 61
```

The code below, creates a plot demonstrating change in positive affect pre- and post-exercise.

```
#set a range of possible X values (i.e., decimal exercise time)
grid_vals <- seq(
  min(EMADat$exerciseHours_std, na.rm = TRUE),
  max(EMADat$exerciseHours_std, na.rm = TRUE),
  length.out = 200
)

#using the final model, Model C, predict PA at each X value
emm <- emmeans(
  PA_modelC,
```

```r
  specs = ~ exerciseHours_std | dummytime,
  at = list(exerciseHours_std = grid_vals)
)

#reverse log-transformed PA values
emm_df <- as.data.frame(emm)
emm_df <- emm_df %>%
  mutate(PA_pred = exp(emmean),
         PA_lower = exp(lower.CL),
         PA_upper = exp(upper.CL))


# Split the data to model pre-exercise and post exercise
emm_before  <- subset(emm_df, dummytime == levels(emm_df$dummytime)[1] & exer
ciseHours_std <= 0)
emm_after   <- subset(emm_df, dummytime == levels(emm_df$dummytime)[2] & exer
ciseHours_std >= 0)

#create a visual plot
ggplot() +
  geom_ribbon(data = emm_before,
              aes(x = exerciseHours_std, ymin = PA_lower, ymax = PA_upper, fi
ll = dummytime),
              alpha = 0.2, color = NA) +
  geom_line(data = emm_before,
            aes(x = exerciseHours_std, y = PA_pred, color = dummytime),
            size = 1.2) +

  geom_ribbon(data = emm_after,
              aes(x = exerciseHours_std, ymin = PA_lower, ymax = PA_upper, fi
ll = dummytime),
              alpha = 0.2, color = NA) +
  geom_line(data = emm_after,
            aes(x = exerciseHours_std, y = PA_pred, color = dummytime),
            size = 1.2) +

  geom_vline(xintercept = 0, linetype = "dashed") +

  scale_color_manual(values = setNames(c("red", "blue"), levels(emm_df$dummyt
ime))) +
  scale_fill_manual(values  = setNames(c("red", "blue"), levels(emm_df$dummyt
ime))) +

  labs(
    x = "Standardized Time in Hours Relative to Longest MVPA Episode",
    y = "Positive Affect",
    color = "Period",
    fill = "Period",
    title = ""
  ) +
```

```
  theme_minimal(base_size = 14) +
  theme(legend.position = "top")

## Warning: Using `size` aesthetic for lines was deprecated in ggplot2 3.4.0.
## ℹ Please use `linewidth` instead.
## This warning is displayed once every 8 hours.
## Call `lifecycle::last_lifecycle_warnings()` to see where this warning was
## generated.
```

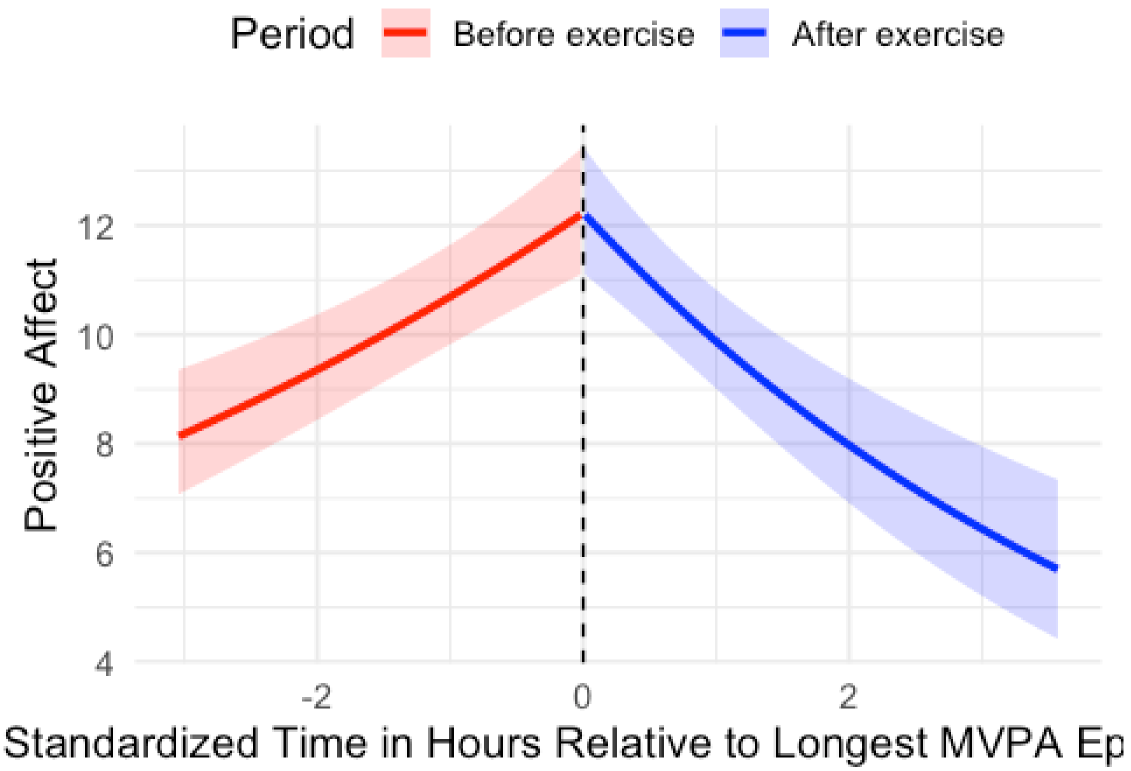


The code below tests calories burned as a moderator to pre- and post- trajectories of PA

```
PA_model_MOD<- lme(
  fixed = log_PosAff ~ 1 + (exerciseHours_std*calories_burned) + ((exerciseHo
urs_std:dummytime)*calories_burned) + (exerciseHours2_std*calories_burned) +
((exerciseHours2_std:dummytime)*calories_burned),
  random = ~ 1 | ID,
  correlation = corCAR1(form = ~ StudyTime | ID),
  data = EMADat,
  na.action = na.omit,
  method = "ML"
)

summary(PA_model_MOD)
```

```
## Linear mixed-effects model fit by maximum likelihood
##   Data: EMADat
##        AIC      BIC    logLik
##   399.5684 467.0276 -186.7842
##
## Random effects:
##  Formula: ~1 | ID
##         (Intercept)  Residual
## StdDev:   0.3143434 0.2679462
##
## Correlation Structure: Continuous AR(1)
##  Formula: ~StudyTime | ID
##  Parameter estimate(s):
##         Phi
## 0.002819198
## Fixed effects:  log_PosAff ~ 1 + (exerciseHours_std * calories_burned) + (
(exerciseHours_std:dummytime) *      calories_burned) + (exerciseHours2_std *
calories_burned) +      ((exerciseHours2_std:dummytime) * calories_burned)
##                                                                     Value
## (Intercept)                                                     2.4425252
## exerciseHours_std                                               0.0513760
## calories_burned                                                 0.0005135
## exerciseHours2_std                                              0.0511548
## exerciseHours_std:calories_burned                               0.0006979
## exerciseHours_std:dummytimeAfter exercise                      -0.2189054
## calories_burned:exerciseHours2_std                              0.0005041
## dummytimeAfter exercise:exerciseHours2_std                      0.0065431
## exerciseHours_std:calories_burned:dummytimeAfter exercise      -0.0010536
## calories_burned:dummytimeAfter exercise:exerciseHours2_std -0.0003758
##                                                             Std.Error   DF
## (Intercept)                                                0.05533375 1255
## exerciseHours_std                                          0.03716953 1255
## calories_burned                                            0.00026037 1255
## exerciseHours2_std                                         0.03581543 1255
## exerciseHours_std:calories_burned                          0.00024110 1255
## exerciseHours_std:dummytimeAfter exercise                  0.09018786 1255
## calories_burned:exerciseHours2_std                         0.00023683 1255
## dummytimeAfter exercise:exerciseHours2_std                 0.03424144 1255
## exerciseHours_std:calories_burned:dummytimeAfter exercise  0.00060824 1255
## calories_burned:dummytimeAfter exercise:exerciseHours2_std 0.00024722 1255
##                                                             t-value p-valu
e
## (Intercept)                                                44.14169  0.000
0
## exerciseHours_std                                           1.38221  0.167
2
## calories_burned                                             1.97200  0.048
8
## exerciseHours2_std                                          1.42829  0.153
5
```

```
## exerciseHours_std:calories_burned                              2.89445  0.003
9
## exerciseHours_std:dummytimeAfter exercise                     -2.42722  0.015
4
## calories_burned:exerciseHours2_std                             2.12870  0.033
5
## dummytimeAfter exercise:exerciseHours2_std                     0.19109  0.848
5
## exerciseHours_std:calories_burned:dummytimeAfter exercise  -1.73222  0.083
5
## calories_burned:dummytimeAfter exercise:exerciseHours2_std -1.52013  0.128
7
##  Correlation:
##                                                            (Intr) exrcH_ c
lrs_b
## exerciseHours_std                                           0.561
## calories_burned                                            -0.520 -0.566
## exerciseHours2_std                                          0.444  0.693 -
0.454
## exerciseHours_std:calories_burned                          -0.427 -0.738
0.791
## exerciseHours_std:dummytimeAfter exercise                  -0.615 -0.909
0.643
## calories_burned:exerciseHours2_std                         -0.335 -0.499
0.621
## dummytimeAfter exercise:exerciseHours2_std                  0.125  0.026 -
0.186
## exerciseHours_std:calories_burned:dummytimeAfter exercise   0.461  0.650 -
0.873
## calories_burned:dummytimeAfter exercise:exerciseHours2_std -0.128 -0.040
0.285
##                                                            exrH2_ exH_:_ e
H_:Ae
## exerciseHours_std
## calories_burned
## exerciseHours2_std
## exerciseHours_std:calories_burned                          -0.511
## exerciseHours_std:dummytimeAfter exercise                  -0.687  0.681
## calories_burned:exerciseHours2_std                         -0.744  0.677
0.499
## dummytimeAfter exercise:exerciseHours2_std                 -0.433 -0.058 -
0.241
## exerciseHours_std:calories_burned:dummytimeAfter exercise   0.487 -0.899 -
0.737
## calories_burned:dummytimeAfter exercise:exerciseHours2_std  0.291  0.106
0.211
##                                                            c_:H2_ dAe:H2 e
H_:_e
## exerciseHours_std
## calories_burned
```

```
## exerciseHours2_std
## exerciseHours_std:calories_burned
## exerciseHours_std:dummytimeAfter exercise
## calories_burned:exerciseHours2_std
## dummytimeAfter exercise:exerciseHours2_std                  0.310
## exerciseHours_std:calories_burned:dummytimeAfter exercise  -0.658  0.233
## calories_burned:dummytimeAfter exercise:exerciseHours2_std -0.361 -0.764 -
0.337
##
## Standardized Within-Group Residuals:
##         Min          Q1         Med          Q3         Max
## -3.66969821 -0.58848615  0.06215655  0.64782559  3.88679831
##
## Number of Observations: 1325
## Number of Groups: 61
```

The code below demonstrates models a-d using ACCELEROMETER exercise time to model negative affect trajectories pre-exercise and post-exercise, including comparisons between models.

MODEL A: NEGATIVE AFFECT

```
#MODEL A: baseline model containing a random effect for participant ID was cr
eated
NA_modelA <- lme(
  fixed = log_NegAff ~ 1,
  random = ~ 1 | ID,
  correlation = corCAR1(form = ~ StudyTime | ID),
  data = EMADat,
  na.action = na.omit,
  method = "ML"
)
summary(NA_modelA)
```

```
## Linear mixed-effects model fit by maximum likelihood
##   Data: EMADat
##        AIC      BIC    logLik
##   262.2733 283.0209 -127.1366
##
## Random effects:
##  Formula: ~1 | ID
##         (Intercept) Residual
## StdDev:   0.2729042 0.256852
##
## Correlation Structure: Continuous AR(1)
##  Formula: ~StudyTime | ID
##  Parameter estimate(s):
##         Phi
## 0.001832901
## Fixed effects:  log_NegAff ~ 1
```

```
##                  Value  Std.Error   DF  t-value p-value
## (Intercept) 2.025665 0.03612687 1261 56.07086       0
##
## Standardized Within-Group Residuals:
##        Min         Q1        Med         Q3        Max
## -3.3192261 -0.6990110 -0.1455217  0.6725808  3.9388616
##
## Number of Observations: 1322
## Number of Groups: 61
```

MODEL B: NEGATIVE AFFECT

```
#MODEL B: two fixed linear slopes were added: 1) hours prior to exercise and
2) hours following exercise
NA_modelB <- lme(
  fixed = log_NegAff ~ 1 + exerciseHours_std + exerciseHours_std:dummytime,
  random = ~ 1 | ID,
  correlation = corCAR1(form = ~ StudyTime | ID),
  data = EMADat,
  na.action = na.omit,
  method = "ML"
)
summary(NA_modelB)
```

```
## Linear mixed-effects model fit by maximum likelihood
##   Data: EMADat
##        AIC      BIC    logLik
##   263.4872 294.6086 -125.7436
##
## Random effects:
##  Formula: ~1 | ID
##         (Intercept)  Residual
## StdDev:   0.2732708 0.2565371
##
## Correlation Structure: Continuous AR(1)
##  Formula: ~StudyTime | ID
##  Parameter estimate(s):
##         Phi
## 0.001802734
## Fixed effects:  log_NegAff ~ 1 + exerciseHours_std + exerciseHours_std:dum
mytime
##                                              Value  Std.Error   DF  t-va
lue
## (Intercept)                              2.0201654 0.03731873 1259 54.13
274
## exerciseHours_std                       -0.0194262 0.01456912 1259 -1.33
338
## exerciseHours_std:dummytimeAfter exercise  0.0132834 0.02287901 1259  0.58
059
##                                           p-value
```

```
## (Intercept)                               0.0000
## exerciseHours_std                         0.1826
## exerciseHours_std:dummytimeAfter exercise  0.5616
##  Correlation:
##                                          (Intr) exrcH_
## exerciseHours_std                         0.207
## exerciseHours_std:dummytimeAfter exercise -0.243 -0.842
##
## Standardized Within-Group Residuals:
##        Min          Q1         Med          Q3         Max
## -3.2639741 -0.7126552 -0.1345223  0.6527646  3.8854841
##
## Number of Observations: 1322
## Number of Groups: 61
```

COMPARE A AND B: NEGATIVE AFFECT

```
#COMPARE A and B#
anova(NA_modelA, NA_modelB)

##           Model df      AIC      BIC    logLik   Test  L.Ratio p-value
## NA_modelA     1  4 262.2733 283.0209 -127.1366
## NA_modelB     2  6 263.4872 294.6086 -125.7436 1 vs 2 2.786037  0.2483
```

MODEL C: NEGATIVE AFFECT

```
#MODEL C: Add two fixed quadratic slopes
NA_modelC <- lme(
  fixed = log_NegAff ~ 1 + exerciseHours_std + exerciseHours_std:dummytime +
exerciseHours2_std + exerciseHours2_std:dummytime,
  random = ~ 1 | ID,
  correlation = corCAR1(form = ~ StudyTime | ID),
  data = EMADat,
  na.action = na.omit,
  method = "ML"
)
summary(NA_modelC)

## Linear mixed-effects model fit by maximum likelihood
##   Data: EMADat
##       AIC      BIC   logLik
##   254.646 296.1412 -119.323
##
## Random effects:
##  Formula: ~1 | ID
##         (Intercept)  Residual
## StdDev:   0.2732937 0.2550853
##
## Correlation Structure: Continuous AR(1)
##  Formula: ~StudyTime | ID
##  Parameter estimate(s):
```

```
##         Phi
## 0.001543185
## Fixed effects:  log_NegAff ~ 1 + exerciseHours_std + exerciseHours_std:dum
mytime +      exerciseHours2_std + exerciseHours2_std:dummytime
##                                                   Value  Std.Error   DF  t-v
alue
## (Intercept)                                    1.9563718 0.04206301 1257 46.5
1051
## exerciseHours_std                             -0.0847470 0.02376747 1257 -3.5
6567
## exerciseHours2_std                            -0.0795541 0.02270926 1257 -3.5
0316
## exerciseHours_std:dummytimeAfter exercise      0.1825152 0.05776733 1257  3.1
5949
## dummytimeAfter exercise:exerciseHours2_std     0.0170664 0.02091587 1257  0.8
1595
##                                               p-value
## (Intercept)                                    0.0000
## exerciseHours_std                              0.0004
## exerciseHours2_std                             0.0005
## exerciseHours_std:dummytimeAfter exercise      0.0016
## dummytimeAfter exercise:exerciseHours2_std     0.4147
##  Correlation:
##                                               (Intr) exrcH_ exrH2_ eH_:Ae
## exerciseHours_std                              0.473
## exerciseHours2_std                             0.372  0.703
## exerciseHours_std:dummytimeAfter exercise     -0.508 -0.914 -0.706
## dummytimeAfter exercise:exerciseHours2_std     0.084  0.025 -0.433 -0.226
##
## Standardized Within-Group Residuals:
##        Min         Q1        Med         Q3        Max
## -3.2233630 -0.6926335 -0.1116408  0.6355782  3.9272320
##
## Number of Observations: 1322
## Number of Groups: 61
```

COMPARE B AND C: NEGATIVE AFFECT

```
#COMPARE B and C#
anova(NA_modelB, NA_modelC)

##           Model df      AIC      BIC    logLik   Test  L.Ratio p-value
## NA_modelB     1  6 263.4872 294.6086 -125.7436
## NA_modelC     2  8 254.6459 296.1412 -119.3230 1 vs 2 12.84128  0.0016
```

MODEL D: NEGATIVE AFFECT

```
NA_modelD <- lme(
  fixed = log_NegAff ~ 1 + exerciseHours_std + exerciseHours_std:dummytime + 
exerciseHours2_std + exerciseHours2_std:dummytime + exerciseHours3_std + exer
ciseHours3_std:dummytime,
```

```
  random = ~ 1 | ID,
  correlation = corCAR1(form = ~ StudyTime | ID),
  data = EMADat,
  na.action = na.omit,
  method = "ML"
)
summary(NA_modelD)
```

```
## Linear mixed-effects model fit by maximum likelihood
##   Data: EMADat
##       AIC      BIC    logLik
##   257.985 309.854 -118.9925
##
## Random effects:
##  Formula: ~1 | ID
##         (Intercept)  Residual
## StdDev:   0.2734543 0.2550277
##
## Correlation Structure: Continuous AR(1)
##  Formula: ~StudyTime | ID
##  Parameter estimate(s):
##         Phi
## 0.001568818
## Fixed effects:  log_NegAff ~ 1 + exerciseHours_std + exerciseHours_std:dum
mytime +      exerciseHours2_std + exerciseHours2_std:dummytime + exerciseHou
rs3_std +      exerciseHours3_std:dummytime
##                                                 Value  Std.Error   DF   t-
value
## (Intercept)                                 1.9899887 0.06725548 1255 29.5
88496
## exerciseHours_std                          -0.0575341 0.04135700 1255 -1.3
91157
## exerciseHours2_std                         -0.0530159 0.05909974 1255 -0.8
97058
## exerciseHours3_std                          0.0078429 0.06649875 1255  0.1
17941
## exerciseHours_std:dummytimeAfter exercise   0.1101578 0.11347631 1255  0.9
70756
## dummytimeAfter exercise:exerciseHours2_std  0.0584823 0.05597981 1255  1.0
44703
## dummytimeAfter exercise:exerciseHours3_std -0.0522037 0.11475758 1255 -0.4
54905
##                                            p-value
## (Intercept)                                 0.0000
## exerciseHours_std                           0.1644
## exerciseHours2_std                          0.3699
## exerciseHours3_std                          0.9061
## exerciseHours_std:dummytimeAfter exercise   0.3319
## dummytimeAfter exercise:exerciseHours2_std  0.2964
## dummytimeAfter exercise:exerciseHours3_std  0.6493
```

```
##  Correlation:
##                                               (Intr) exrcH_ exrH2_ exrH3_ eH_
:Ae
## exerciseHours_std                              0.619
## exerciseHours2_std                             0.784  0.525
## exerciseHours3_std                             0.562  0.012  0.811
## exerciseHours_std:dummytimeAfter exercise     -0.813 -0.868 -0.832 -0.461
## dummytimeAfter exercise:exerciseHours2_std     0.662  0.727  0.567  0.299 -0.
819
## dummytimeAfter exercise:exerciseHours3_std -0.741 -0.366 -0.922 -0.901  0.
731
##                                               dAe:H2
## exerciseHours_std
## exerciseHours2_std
## exerciseHours3_std
## exerciseHours_std:dummytimeAfter exercise
## dummytimeAfter exercise:exerciseHours2_std
## dummytimeAfter exercise:exerciseHours3_std -0.651
##
## Standardized Within-Group Residuals:
##        Min         Q1        Med         Q3        Max
## -3.2170074 -0.7002285 -0.1093427  0.6400770  3.9474342
##
## Number of Observations: 1322
## Number of Groups: 61
```

COMPARAE C AND D: NEGATIVE AFFECT

```
#COMPARE C and D#
anova(NA_modelC, NA_modelD)
```

```
##           Model df     AIC      BIC    logLik   Test   L.Ratio p-value
## NA_modelC     1  8 254.646 296.1412 -119.3230
## NA_modelD     2 10 257.985 309.8540 -118.9925 1 vs 2 0.6609908  0.7186
```
